# Impact of public and private sector COVID-19 diagnostics and treatments on US healthcare resource utilization

**DOI:** 10.1101/2020.11.09.20228452

**Authors:** Daniel Sheinson, William Wong, Caroline Solon, Mindy Cheng, David Elsea, Yang Meng, Anuj Shah

## Abstract

**Introduction:** Coronavirus disease 2019 (COVID-19) has imposed a considerable burden on the United States (US) health system, with particular concern over healthcare capacity constraints.

**Methods:** We modeled the impact of public and private sector contributions to developing diagnostic testing and treatments on COVID-19-related healthcare resource use.

**Results:** We estimated that public sector contributions lead to ≥30% reductions in COVID-19-related healthcare resource utilization. Private sector contributions to expanded diagnostic testing and treatments lead to further reductions in mortality (−44%), intensive care unit (ICU) and non-ICU hospital beds (−30% and −28%, respectively), and ventilator use (−29%). The combination of lower diagnostic test sensitivity and proportions of patients self-isolating may exacerbate case numbers, and policies that encourage self-isolating should be considered.

**Conclusion:** While mechanisms exist to facilitate research, development, and patient access to diagnostic testing, future policies should focus on ensuring equitable patient access to both diagnostic testing and treatments which, in turn, will alleviate COVID-19-related resource constraints.

## Introduction

As of September 23, 2020, over 30 million coronavirus disease 2019 (COVID-19) cases are estimated to have occurred worldwide, resulting in more than 900,000 deaths[1] - of these, the United States (US) is estimated to account for over 6 million cases and more than 200,000 deaths. Despite constantly evolving data as the pandemic progresses, early estimates suggest approximately 20% of COVID-19 infected patients require hospitalization[2]. Of adults hospitalized in the US, estimates suggest 32% required intensive care unit (ICU) admission and 19% required invasive mechanical ventilation[3], representing a considerable burden to the health system.

Health system capacity has been a widely reported concern in the US since the onset of the pandemic, with many studies highlighting an insufficient number of hospital beds, ICU beds, and mechanical ventilators to address the additional needs anticipated with COVID-19[4-6]. The dramatic increase in need for health care resources by COVID-19 patients with severe manifestations that require more intensive treatment risks overwhelming hospital systems, in terms of both staff and space to treat patients[5]. For example, the New York Public Health System had a pre-pandemic capacity of 300 ICU beds, yet at the peak of the COVID-19 surge were caring for 1,000 ICU patients[7]. This burden is further compounded by the longer lengths of stay (LOS) required by COVID-19 patients requiring respiratory support, with estimates of ICU ventilator use ranging from 18-23 days vs. approximately 3-8 days during non-pandemic times[8-10]. In turn, capacity constraints - particularly in the ICU setting - may result in increased mortality rates[11-14].

Early models focused on estimating the impact of COVID-19 on hospital system capacity metrics, including admissions, ICU admissions, and mechanical ventilator use[6, 15]. However, since then, due to the complex challenges of COVID-19, both the public and private sectors have contributed significantly to increasing diagnostic testing capacity, beyond public health laboratories and individual laboratory developed tests (LDTs), and studying treatments to address COVID-19. For example, in April 2020, the Accelerating COVID-19 Therapeutic Interventions and Vaccines (ACTIV) partnership was formed between members of the public and private sectors to develop a coordinated research strategy to help accelerate the development and distribution of COVID-19 therapeutics and vaccines[16]. Given the rapid development of innovations to address COVID-19, this presents an opportunity to develop models focusing specifically on the impacts of diagnostic testing and therapeutic intervention that have not been accounted for in previous models.

Despite both public and private sectors accelerating diagnostic testing and treatments through Food & Drug Administration (FDA) emergency use authorizations (EUAs), there remains a large degree of uncertainty regarding the impact of COVID-19 testing and treatment at reducing time to recovery, ICU or ventilator use, and/or mortality. Therefore, the objective of this research was to develop a model to estimate the impact to date of public and private sector contributions to developing effective COVID-19 treatment and nucleic acid-based (molecular) diagnostic testing on resource use in the hospital (including hospital beds, ICU beds, and mechanical ventilators) and overall population impacts (including mortality and cumulative incidence of disease) at the US national level during the pandemic.

## Methods

### Model Structure

An age-stratified compartmental model of disease transmission and healthcare resource utilization across the entire U.S. was adapted from Moghadas *et al*.[6] and further developed by incorporating the effects of diagnostic testing among infected individuals and novel treatment in the hospital setting. While the transmission rates in the U.S. have been shown to vary by geographic region, a model at this level would require region-specific details around testing and treatment capacities which are not publicly available. Hence, we aimed to estimate disease spread and resource use in aggregate across the country (see Discussion section for limitations). In our model, individuals start as uninfected but susceptible to disease and potentially become exposed to infection according to a time-varying transmission rate. Once exposed, individuals develop asymptomatic, mild, or severe disease. A subset of symptomatic individuals self-isolate, thereby reducing their contact rate with others in the population and the overall transmission of infection. Asymptomatic and mildly symptomatic individuals were assumed to recover from infection while a proportion of severely symptomatic individuals incur different levels of hospital resource utilization in the form of general ward (non-ICU), ICU admission, or ICU admission with mechanical ventilation (see Figure 1). Hospitalized patients either recover or die according to separate rates of recovery and mortality for each level of care. Recovered individuals were assumed to be immune and not infect others. Four age categories (0-19, 20-49, 50-64, 65+) were modeled throughout, allowing for age-specific transition probabilities of developing severe illness, hospitalization, and ICU admission. The model was implemented in Microsoft Excel using a Markov structure with 1-day cycles and was initialized by assuming 1 exposed individual in each age group at day 0 corresponding to January 21, 2020.

**Figure 1.**
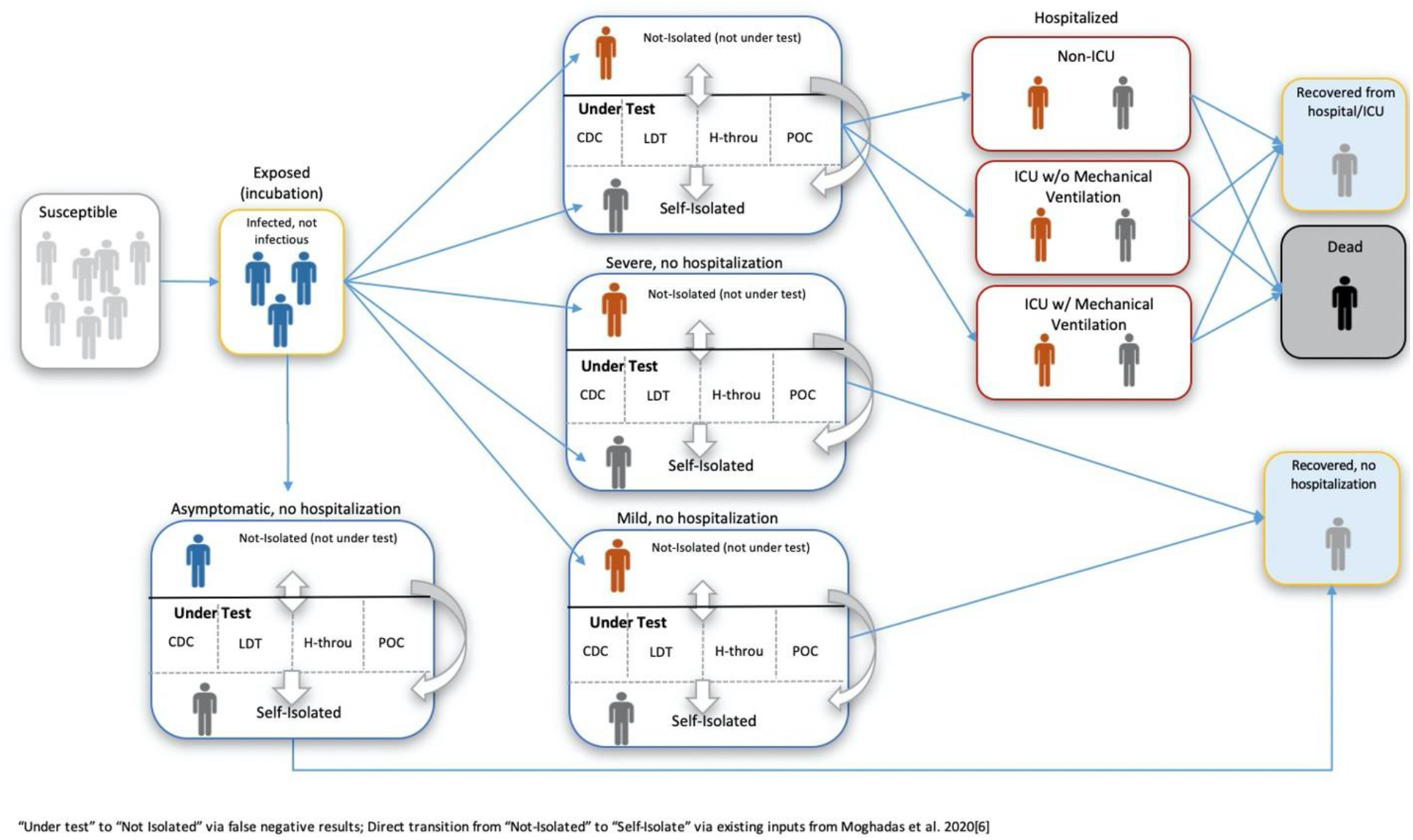
Compartmental model structure

Diagnostic testing was incorporated into the model by assuming that some infected individuals undergo testing, upon which a positive test result leads to self-isolation of that individual. Treatment was incorporated into the model through specific inputs affecting the rates of hospital recovery, mortality, and mechanical ventilation usage. Additional model input parameters control the size and age distribution of the population, rates of transmission and recovery, rates of testing and time to results, symptom onset and severity, rates of self-isolation, levels of hospitalization, mortality, and testing capacity.

### Model Calibration and Data Sources

Published literature, government, and non-government sources on COVID-19 were gathered to inform input parameter values and provide prior information for calibrating the model to observed data from The COVID Tracking Project[17] (see appendix in electronic supplementary materials for detailed methodology). Model parameters related to disease transmission, symptoms, self-isolation, and testing were obtained mainly from early publications on the spread of the disease in China[18, 19] and input into the model directly (see appendix Tables S1 and S2 in the electronic supplementary materials for details). Model parameters related to hospitalization, including hospitalization rate, proportion admitted by care setting, and rates of death and recovery were obtained mainly from US-specific sources and informed prior distributions for calibrating those parameters to the observed data (see appendix Table S4). In addition, time-varying rates of transmission and testing were incorporated into the model and – along with the aforementioned hospitalization parameters – calibrated to the observed number of daily positive tests, patients in the hospital, patients in the ICU, patients on mechanical ventilation, and cumulative deaths using a sequential estimation technique called the kernel density particle filter[20]. Model calibration was implemented using R version 3.5.3[21], and the programming code is available on Github (https://github.com/Roche/covid-hcru-model).

### Model Scenarios

Once the model was calibrated, we simulated five scenarios to compare expected transmission, hospital resource use, and mortality: 1) a reference scenario reflecting no public or private sector contributions to diagnostic testing or treatment; 2) a public sector only scenario reflecting only diagnostic testing and treatment contributions from the public sector; 3) a scenario with public and private sector contributions in developing effective treatment only; 4) a scenario with public and private sector contributions in developing effective diagnostic testing only; 5) a scenario with public and private sector contributions in developing both effective treatment and testing. Each scenario was generated using modified input parameters reflecting the assumed level of testing and treatment in the population. Model results for each scenario are reported for a time period when private sector diagnostic tests and effective novel treatments were both available via FDA EUAs. This was defined to be from June 1, 2020 to the latest date where COVID-19 tracking data were available at the time of model calibration (August 21, 2020).

### Molecular diagnostic testing

Test systems considered in the model included commercial high-throughput molecular assays (HT; as defined in the Centers for Medicare and Medicaid Services [CMS] Ruling 2020-01-R as of April 14, 2020[22]) and molecular point-of-care assays (POC), which received FDA EUA by June 1, 2020; other commercial tests (Other); a test developed by the Centers for Disease Control and Prevention (CDC); and independent LDTs. In the public sector only scenario, it was assumed that only LDTs and the CDC test for SARS-CoV-2 diagnosis would be available (i.e. no HT or POC tests available). In scenarios that included the effects of test systems developed through the private sector, the capacity of HT, POC, and other commercial tests were set according to market projections based on publicly available data reported by major test manufacturers. Market share assumptions for the different test systems were based on survey data[23] on their relative use in laboratories.

### Treatment efficacy

Given its established availability prior to the pandemic and the RECOVERY study[24], which demonstrated its clinical benefit for COVID-19, and being funded primarily via ex U.S. sources, dexamethasone was assumed to be an available treatment option in the public sector only scenario. In the scenarios including treatments developed from the private sector, treatment effects on hospital recovery and mortality rates were based on data for remdesivir, since it was the only new treatment that was issued an EUA by the FDA for COVID-19[25] at the time of model calibration. Given the recent data suggesting that there may be some uncertainty surrounding the efficacy of remdesivir [26, 27], we conducted a number of sensitivity analyses (see ‘Scenario and sensitivity analyses’ below). To be conservative with respect to some eligible patients not receiving treatment or discontinuing their treatment early, it was assumed that 50% of eligible patients were treated.

### Scenario and sensitivity analyses

Additional scenario analyses were conducted to explore the inclusion of additional treatment effects on mortality and reduced use of mechanical ventilation. Given the potential uncertainty in the treatment effects of remdesivir[26, 27], a scenario analysis was also conducted for which the private sector effects were due to testing only (i.e. remdesivir treatment effects were excluded). One-way and two-way sensitivity analyses were conducted to quantify the impacts of uncertainty associated with individual model parameters, including the market share of HT testing, the time at which public-private testing and treatment were available, and the impact of test sensitivity versus the proportion of patients that self-isolate while awaiting their test results.

## Study results

Calibration of model parameters resulted in estimated resource utilization and cumulative mortality that tracked with observed data from The COVID Tracking Project (Figure 2). In the absence of public and private sector contributions (reference scenario), we estimated a cumulative incidence of 17,952,264 COVID-19 cases, a cumulative mortality of 315,230 deaths, peak (i.e. maximum) non-ICU hospital beds needed of 153,698, peak ICU beds needed of 37,245, and peak ventilator use of 15,338 ventilators from June 1st, 2020 to August 21, 2020 in the US (Table 1; Figure 3a-d). In the public sector only scenario, which assumed availability of only the CDC test and LDTs without novel effective treatments attributed to the private sector, we estimated ≧30% reductions in all outcomes measured relative to the reference scenario, with the largest effect on cumulative mortality (−40.2%). The addition of private sector contributions to diagnostic testing capacity and effective treatment options were associated with additional reductions (relative to the public sector only scenario) across all outcomes, with the greatest effect on cumulative mortality (−44.0%) and reductions on other outcomes ranging from −15.2% to −29.9% (Table 1).

**Table 1.**
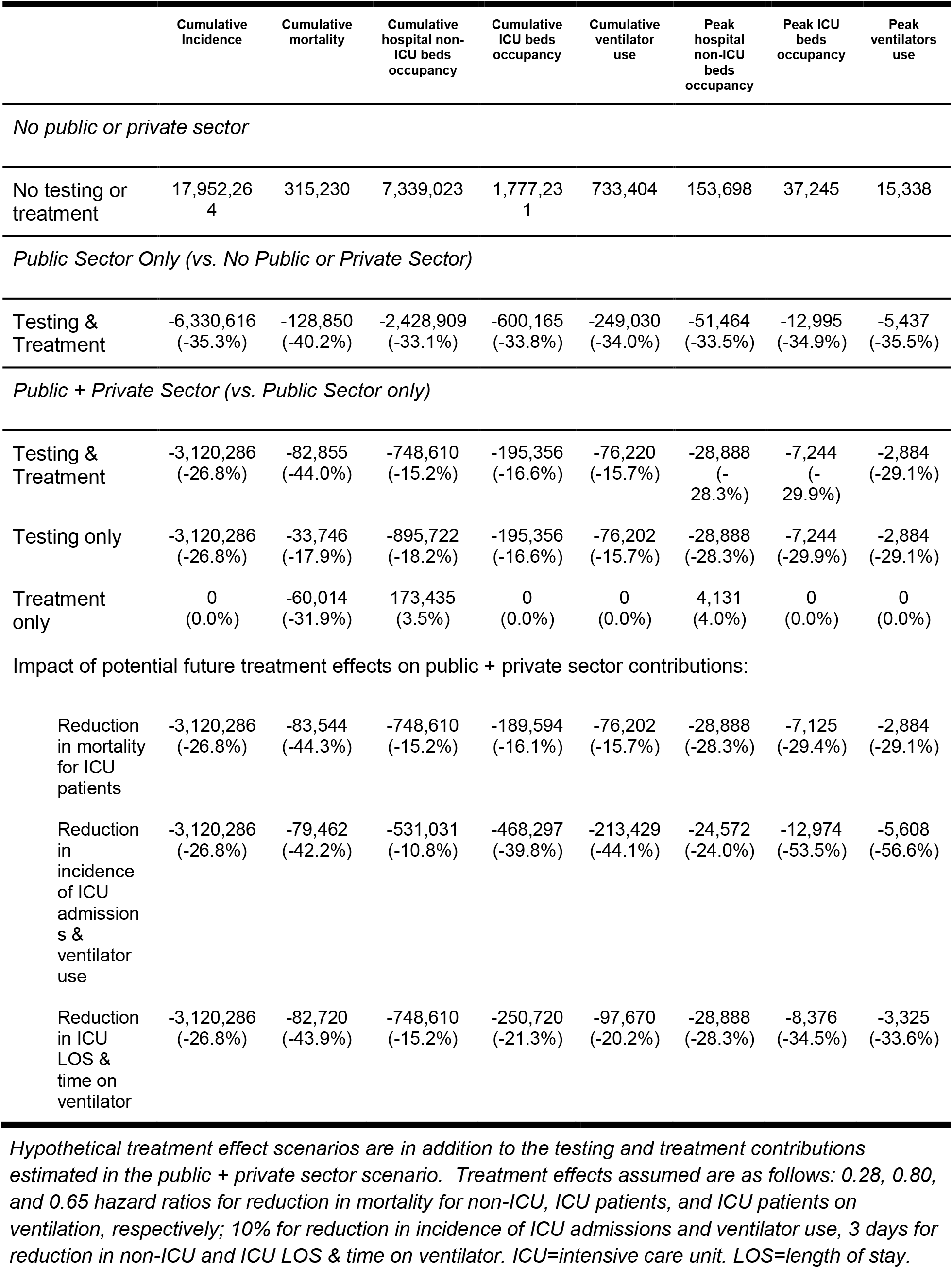
Estimated effect of public and private sector contributions to the reduction in the incidence and mortality of COVID-19 and improvements in health care resource utilization between June 1 to August 21, 2020

|  | Cumulative Incidence | Cumulative mortality | Cumulative hospital non-ICU beds occupancy | Cumulative ICU beds occupancy | Cumulative ventilator use | Peak hospital non-ICU beds occupancy | Peak ICU beds occupancy | Peak ventilators use |
| --- | --- | --- | --- | --- | --- | --- | --- | --- |
| *No public or private sector* |  |  |  |  |  |  |  |  |
| No testing or treatment | 17,952,264 | 315,230 | 7,339,023 | 1,777,231 | 733,404 | 153,698 | 37,245 | 15,338 |
| *Public Sector Only (vs. No Public or Private Sector)* |  |  |  |  |  |  |  |  |
| Testing & Treatment | -6,330,616 (-35.3%) | -128,850 (-40.2%) | -2,428,909 (-33.1%) | -600,165 (-33.8%) | -249,030 (-34.0%) | -51,464 (-33.5%) | -12,995 (-34.9%) | -5,437 (-35.5%) |
| *Public + Private Sector (vs. Public Sector only)* |  |  |  |  |  |  |  |  |
| Testing & Treatment | -3,120,286 (-26.8%) | -82,855 (-44.0%) | -748,610 (-15.2%) | -195,356 (-16.6%) | -76,220 (-15.7%) | -28,888 (-28.3%) | -7,244 (-29.9%) | -2,884 (-29.1%) |
| Testing only | -3,120,286 (-26.8%) | -33,746 (-17.9%) | -895,722 (-18.2%) | -195,356 (-16.6%) | -76,202 (-15.7%) | -28,888 (-28.3%) | -7,244 (-29.9%) | -2,884 (-29.1%) |
| Treatment only | 0 (0.0%) | -60,014 (-31.9%) | 173,435 (3.5%) | 0 (0.0%) | 0 (0.0%) | 4,131 (4.0%) | 0 (0.0%) | 0 (0.0%) |
| Impact of potential future treatment effects on public + private sector contributions: |  |  |  |  |  |  |  |  |
| Reduction in mortality for ICU patients | -3,120,286 (-26.8%) | -83,544 (-44.3%) | -748,610 (-15.2%) | -189,594 (-16.1%) | -76,202 (-15.7%) | -28,888 (-28.3%) | -7,125 (-29.4%) | -2,884 (-29.1%) |
| Reduction in incidence of ICU admissions & ventilator use | -3,120,286 (-26.8%) | -79,462 (-42.2%) | -531,031 (-10.8%) | -468,297 (-39.8%) | -213,429 (-44.1%) | -24,572 (-24.0%) | -12,974 (-53.5%) | -5,608 (-56.6%) |
| Reduction in ICU LOS & time on ventilator | -3,120,286 (-26.8%) | -82,720 (-43.9%) | -748,610 (-15.2%) | -250,720 (-21.3%) | -97,670 (-20.2%) | -28,888 (-28.3%) | -8,376 (-34.5%) | -3,325 (-33.6%) |

Hypothetical treatment effect scenarios are in addition to the testing and treatment contributions estimated in the public + private sector scenario. Treatment effects assumed are as follows: 0.28, 0.80, and 0.65 hazard ratios for reduction in mortality for non-ICU, ICU patients, and ICU patients on ventilation, respectively; 10% for reduction in incidence of ICU admissions and ventilator use, 3 days for reduction in non-ICU and ICU LOS & time on ventilator. ICU=intensive care unit. LOS=length of stay.

**Figure 2.**
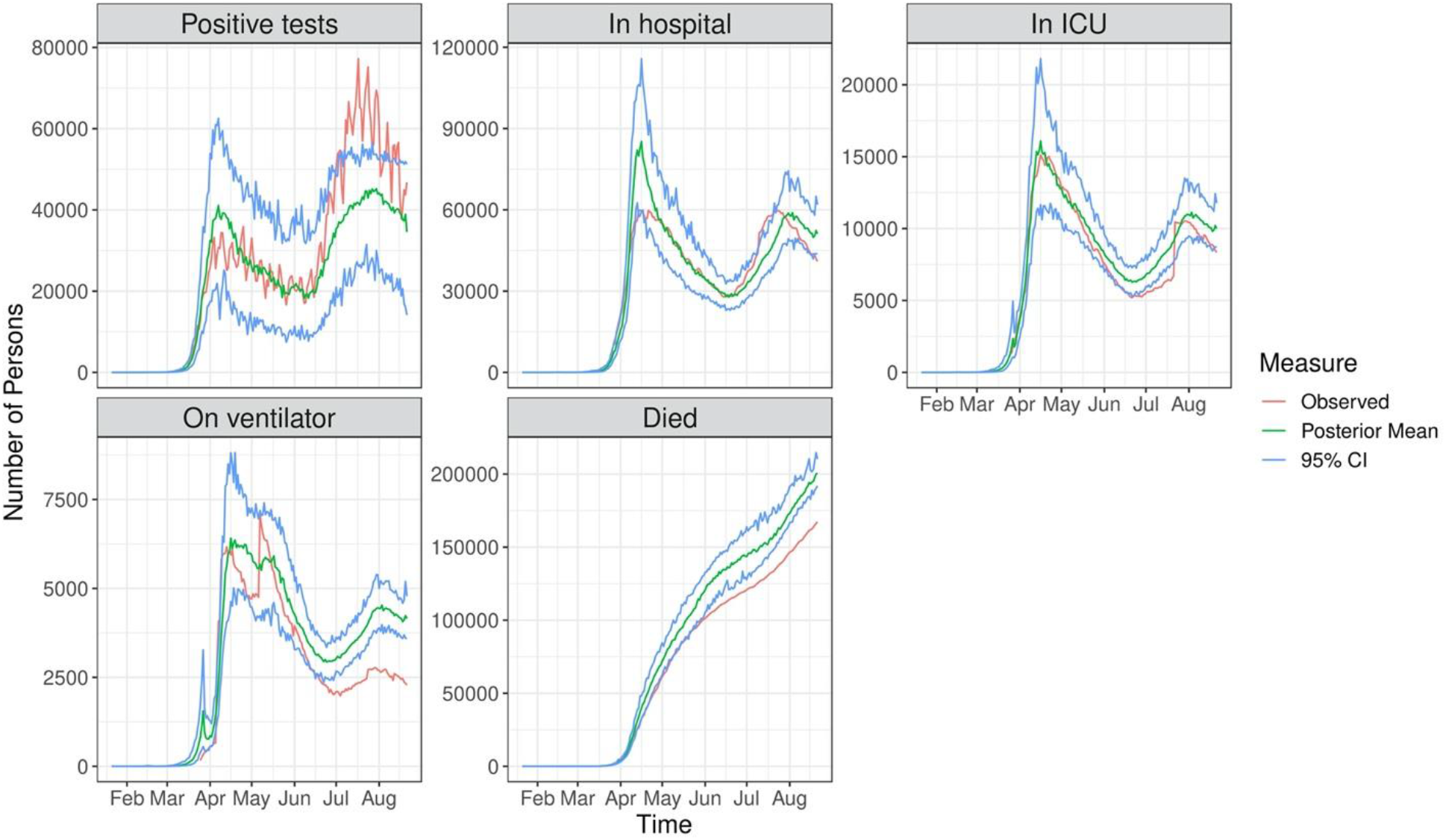
Estimated resource utilization (posterior means) over time and 95% credible intervals from calibrated model alongside observed data from The COVID Tracking Project.

**Figure 3.**
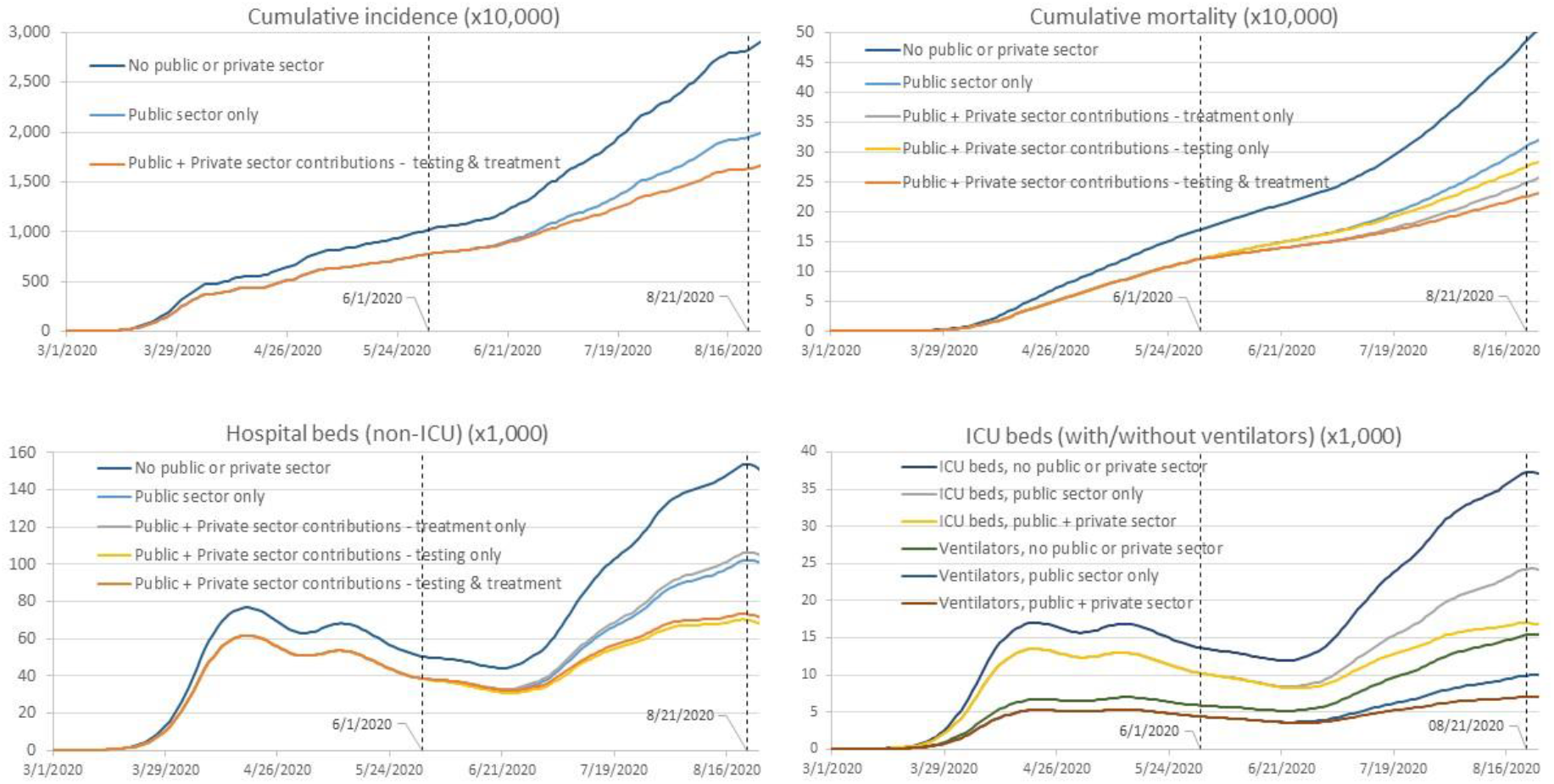
Public and private sector contributions for COVID-19 incidence (a) and mortality (b) and hospital resource use over time (c, d) 6/1/2020 is the assumed start date of the private sector contributions to diagnostic testing and novel effective treatments. 8/21/20 is the end date of the results reporting period. ICU=Intensive care unit.

Examining the individual effects of the private sector contributions allows us to infer the relative contributions of expanded diagnostic testing versus novel treatment that contribute to reductions in individual outcomes. The public + private sector scenario assumed novel treatment from the private sector provided only a mortality benefit and reduced LOS among non-ICU hospitalized patients. Therefore, the scenario with novel effective treatment alone only affected cumulative mortality (60,014 fewer deaths; - 31.9%), cumulative non-ICU hospital occupancy (173,435 more beds; +3.5%), and peak non-ICU hospital occupancy (4,131 more beds; +4.0%). In this case, the mortality benefit leads to greater non-ICU hospital resource use due to longer LOS among non-ICU patients that recover compared to those that die.

The individual effect of private sector expansion of commercial diagnostic testing capacity alone (i.e. excluding any private sector treatment effects) affected all outcomes. This was due to the increased number of true positive test results leading to greater self-isolation and reduced transmission, which has downstream effects on hospital resource use and mortality. In this scenario, expanded testing contributed to 3,120,286 fewer cases (−26.8%), 33,746 fewer deaths (−17.9%), 31,883 fewer non-ICU beds at peak occupancy (−31.2%), 7,244 fewer ICU beds at peak occupancy (−29.9%), and 2,884 fewer ICU patients on mechanical ventilation at peak usage (−29.2%). Taken together, we can infer that for the combined effects, expanded diagnostic testing was the larger contributor to reducing non-ICU hospital bed occupancy while effective novel treatment had a greater contribution to reducing mortality.

Scenario analyses examining the various effects of treatment, as a proxy for potential new treatments which may have different effects than those currently marketed, demonstrated a potential reduction in outcomes not previously impacted by current treatment via public and private sector contributions (Table 1). When treatment-related reductions in ICU admissions and ventilator use were applied, the estimated reduction in peak ICU beds increased 1.8-fold (−53.5% vs. −29.9%) and cumulative ICU beds increased 2.4-fold (−39.8% vs. −16.6%). Similarly, assuming reductions in ICU LOS and time on ventilator further reduced peak and cumulative ICU beds, but to a lesser extent (−34.5% vs. −29.9%, −21.3% vs. −16.6%).

In sensitivity analyses, changing the date at which expanded diagnostic testing and novel treatment were assumed to be available from June 1st to May 1st (as a proxy for when remdesivir was first available via EUA) resulted in further reductions in resource utilization, mortality, and incidence of COVID-19 cases (range: −11.8% to −16.1%; Table 2). Among the diagnostic testing parameters varied, the model was highly sensitive to assumptions around time to test result and LDT sensitivity. Increasing time to test result by just 1 day (from 3 to 4 days) increased the cumulative incidence of cases by +27.9% and resources utilized by +17.9-32.6%. Similarly, varying LDT sensitivity from 100% to as low as 60% resulted in large changes in the incidence of COVID-19 cases (−4.9% to +71.3% cases) and resource use (ranging from −5.3% to +77.3%). Shifting 20% of the share of available testing systems toward more HT tests resulted in changes in all outcomes ranging from −0.8% to −2.7%. Patient compliance with self-isolation while awaiting test results was also found to have a significant effect on the cumulative incidence of cases and accentuated the effects of lower testing accuracy (Figure 4). For example, the change in cumulative incidence of COVID-19 cases for diagnostic test sensitivities ranging from 100% to 85% was between −10.2% and +50.0% for the assumed level of 70% self-isolation while awaiting test results. However, when self-isolation while awaiting test results was decreased to 50%, change in cumulative incidence of COVID-19 cases ranged from +219.8% to +340.3%. Model sensitivity to other clinical and behavioral factors are shown in Table 2.

**Table 2.** Impact of model assumptions and parameters on incidence and mortality of COVID-19 and health care resource utilization

|  | (L, H) | Relative change vs. public+private sector contribution scenario |  |  |  |  |  |  |  |
| --- | --- | --- | --- | --- | --- | --- | --- | --- | --- |
|  |  | Cumulative incidence | Cumulative mortality | Cumulative hospital non-ICU beds occupancy | Cumulative ICU beds occupancy | Cumulative ICU beds (ventilators) occupancy | Peak hospital non-ICU beds occupancy | Peak ICU beds occupancy | Peak ICU beds (ventilators) occupancy |
| **Public + private sector contribution - treatment and testing** |  | **8,501,362** | **105,525** | **4,162,323** | **981,710** | **408,155** | **73,346** | **17,006** | **7,016** |
| **Private sector contribution starting date** |  |  |  |  |  |  |  |  |  |
| Public + Private sector contribution starting date May 1<sup>a</sup> |  | -15.4% | -12.5% | -12.6% | -12.3% | -11.8% | -16.1% | -15.9% | -15.7% |
| **Testing scenarios** |  |  |  |  |  |  |  |  |  |
| Increase in HT testing by 20%<sup>b</sup> |  | -2.0% | -0.9% | -0.9% | -0.8% | -0.8% | -2.7% | -2.6% | -2.4% |
| % testing performed for asymptomatic patients | (0%, 20%) | (-3.4%, 3.3%) | (-2.9%, 2.8%) | (-2.9%, 2.8%) | (-2.8%, 2.6%) | (-2.7%, 2.6%) | (-3.9%, 3.7%) | (-3.8%, 3.6%) | (-3.7%, 3.6%) |
| LDT test sensitivity | (60%, 100%) | (71.3%, -4.9%) | (61.3%, -4.1%) | (61.7%, -4.1%) | (59.0%, -4.0%) | (57.6%, -3.9%) | (77.3%, -5.3%) | (75.6%, -5.2%) | (74.6%, -5.1%) |
| Time to test result - days | (2, 4) | (-22.1%, 27.9%) | (-18.5%, 19.8%) | (-18.7%, 20.0%) | (-17.9%, 18.7%) | (-17.4%, 17.9%) | (-24.2%, 32.6%) | (-23.5%, 31.2%) | (-23.2%, 30.4%) |
| **Clinical scenarios** |  |  |  |  |  |  |  |  |  |
| Time from symptom onset to test for severe/critical patients - days | (0.5, 2) | (-33.2%, 37.4%) | (-31.2%, 36.5%) | (-31.3%, 36.6%) | (-30.7%, 35.8%) | (-30.4%, 35.4%) | (-34.3%, 38.3%) | (-34.0%, 38.0%) | (-33.8%, 37.9%) |
| Time from symptom onset to test for mild patients - days | (1, 3) | (-26.1%, 10.1%) | (-24.3%, 9.5%) | (-24.3%, 9.6%) | (-23.8%, 9.3%) | (-23.5%, 9.2%) | (-27.3%, 10.6%) | (-27.0%, 10.5%) | (-26.8%, 10.4%) |
| % of infected individuals that are asymptomatic | (10%, 25%) | (16.7%, -13.0%) | (24.9%, -18.7%) | (25.1%, -18.8%) | (24.3%, -18.4%) | (23.9%, -18.1%) | (30.7%, -22.1%) | (29.9%, -21.7%) | (29.5%, -21.5%) |
| **Behavioral factors** |  |  |  |  |  |  |  |  |  |
| % of severe cases self-isolating immediately upon symptom onset | (0%, 10%) | (9.9%, -9.2%) | (8.7%, -8.0%) | (8.7%, -8.1%) | (8.4%, -7.8%) | (8.3%, -7.7%) | (11.1%, -10.1%) | (10.8%, -9.9%) | (10.6%, -9.7%) |
| % severe cases self-isolating after symptom onset | (70%, 90%) | (5.1%, -4.5%) | (4.8%, -4.2%) | (4.8%, -4.3%) | (4.7%, -4.2%) | (4.7%, -4.1%) | (5.3%, -4.7%) | (5.2%, -4.6%) | (5.2%, -4.6%) |
| % mild cases self-isolating after symptom onset | (0%, 10%) | (13.1%, -11.8%) | (12.3%, -11.0%) | (12.4%, -11.0%) | (12.1%, -10.8%) | (11.9%, -10.6%) | (13.7%, -12.3%) | (13.5%, -12.2%) | (13.4%, -12.1%) |

<sup>a</sup>Assumes private sector contributions to treatment and testing starts on May 1, 2020, and the result collection period is the same as base case June 1 to August 21. <sup>b</sup>The number of HT machines are increased by 20% and the number of LDT is reduced by the same absolute amount so that the total
number of systems are the same. H=high parameter estimate. HT=high throughput. ICU=intensive care unit. L=low parameter estimate. LB=lower bound. LDT=Laboratory Developed Test.

**Figure 4.**
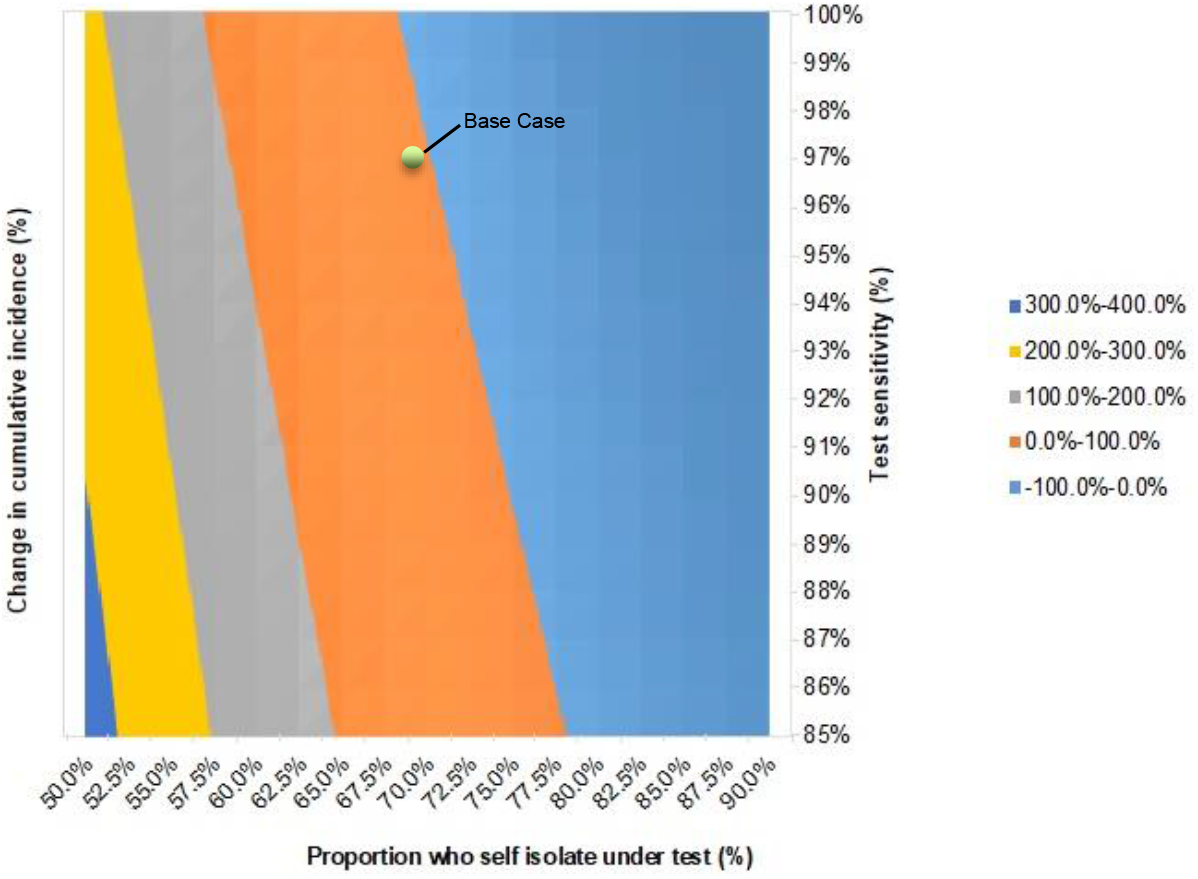
The relationship between test sensitivity and self-isolation while awaiting test results on cumulative incidence of COVID-19 cases Results displayed are the percentage change in cumulative incidence relative to the scenario assuming public + private sector contributions to diagnostic testing and novel treatment.

## Discussion

To our knowledge, this is the first study to comprehensively estimate the contributions of the public and private sectors, via development of molecular diagnostic testing and treatments, in addressing COVID-19-related health system resource constraints in the US. We found that the effect of public sector contributions likely had a significant effect on altering the trajectories of cases, mortality, and resource utilization. Furthermore, EUAs expanding and expediting the availability of commercial diagnostic tests and therapeutics to treat COVID-19 likely had a significant impact on providing further reductions in the incidence of COVID-19, peak hospital capacities, ventilator use, and mortality. This finding is consistent despite uncertainty in private sector treatment effects, indicating diagnostics alone may have a role in reducing heath resource utilization, although the combination of both testing and treatments together produced a greater effect than either considered alone. While this model focuses on the cases and resource use from a US national perspective, regional differences in COVID-19 have been well documented. Since the availability of testing and treatment resources may vary regionally and not always align with the number of cases present in a particular geography, our model provides an optimistic scenario and highlights the importance of ensuring efficient and equitable distribution of treatments and diagnostics.

While this study focused on health care resource utilization, it is also important to consider the economic implications of the reduction in resources. Estimates for the COVID-19 health care cost per day to hospitals ranges from $2,303 (non-ICU) to $3,449 (ICU with ventilation)[28]. With hospitals facing substantial financial burden due to COVID-19, any diagnostic test or treatment which can reduce hospitalizations and LOS may provide significant cost savings. While estimating costs of interventions and hospitalizations were beyond the scope of this study, it should be noted that the costs of a course of treatment for the only guideline recommended COVID-19 treatments during this study period (remdesivir and dexamethasone)[29-31] are less than, or similar to, a single day in a hospital. The potential economic benefits of these interventions due to health system capacity impacts should be considered when evaluating the value of a technology (diagnostic or treatment).

Based on our findings, public and private sector contributions have played an important role in addressing the pandemic; however, critical needs remain. Despite the increase in diagnostic testing capacity since the beginning of the pandemic, laboratory backlogs have been observed[32]. As estimated in this study, time to test result is an important factor in reducing laboratory backlog and impacts the infection curve and hospitalizations. As a result of the pressing need to increase laboratory testing capacity, other factors, including test sensitivity have been less scrutinized. Our findings highlight that test sensitivity is an important factor to consider, as inaccurate test results pose risk to further viral transmission. In particular, the real-world clinical validity of available test options is largely unknown, and our results demonstrate that tests with lower sensitivity (i.e. increased false negative results) may significantly contribute toward disease transmission and health care resource utilization. Based on this, it is important to consider test system capacity, time to test result, and test sensitivity when evaluating the potential effectiveness of different testing strategies. Additionally, behavioral factors, such as patient compliance with self-isolation, are crucial toward minimizing viral transmission. We found the combination of poor patient self-isolation behaviors and lower test sensitivity can accentuate the impact of false negatives on disease transmission and subsequent healthcare system resource utilization. Therefore, when considering trade-offs between test sensitivity and time to test result, patient compliance with self-isolation behaviors is an important parameter to understand. Consequently, policies that support and enable people to self-isolate without penalties or risks (e.g. loss of employment, school, etc.) are important.

Effective treatments are equally important given the complementary effects of diagnostic testing on health care resource use and patient outcomes in COVID-19. Recent therapies issued EUA have involved both private sector contributions (e.g. remdesivir), as well as public funding (e.g. dexamethasone, convalescent plasma). In this study, we found that included treatments (remdesivir, dexamethasone) only impacted mortality and non-ICU hospital resource use. That said, with research and development (R&D) occurring at a record pace, there are over 300 therapies under investigation for COVID-19[33], and these new potential treatments may impact different aspects of health care resource use (i.e. prevention of hospitalization, reduction in ventilator use, etc.) which can further substantially reduce capacity constraints. This may be particularly important when considering the individual effects of treatments, as those which reduce mortality may increase hospital stays due to the prolonged survival effect, as observed in our study where the effect of diagnostic testing alone led to a greater reduction in non-ICU beds than with testing and treatment. Therefore, a potentially optimal scenario may be a combination of available novel treatments that balance the reduction in mortality and LOS outcomes. Furthermore, while COVID-19 vaccines were not modeled in this present study, it could be expected that they would have a hugely beneficial impact on health system resource constraints and patient outcomes via reductions in infectivity and subsequent hospitalizations and mortality. That said, production and distribution of any vaccine at scale may be challenging; thus, development of novel treatments may have similar or even greater importance than vaccine development. However, unlike the rapid progress in innovation of diagnostic testing, developing therapeutics has been more challenging, with many failed trials highlighting the difficulty of finding success in R&D. Policies which continue to facilitate R&D are critical for the ability to develop innovative approaches to addressing the COVID-19 pandemic.

In addition to policies which facilitate R&D and allow recent innovations to be quickly available, those which enable patient access to these innovations are equally important. For example, the Families First Coronavirus Response Act (FFCRA) ensured that most patients would not incur out of pocket costs for COVID-19 diagnostic testing[34]. The Coronavirus Aid, Relief and Economic Security (CARES) Act amended the FFCRA to further allow coverage of testing without cost-sharing, including those receiving testing out of network[34]. The significant impact diagnostic testing may have on multiple aspects of health care resource utilization, as observed in this study, highlights the importance of these policies which have facilitated patient access to testing resources. While beyond the scope of this analysis, the absence of these policies may have resulted in diminished reductions in incident cases and resource use due to the public and private sector contributions modeled in this study; however, future research would be needed to confirm this hypothesis. Additionally, despite the enactment of these policies, there may still be subgroups of patients who are not covered by FFCRA or CARES, such as the uninsured. Given the rapid developments and increasing availability of various testing technologies (e.g. antibody, antigen, multiplex molecular tests, etc.), additional policies that clarify coverage requirements and facilitate patient access to COVID-19 testing across the varying technologies should be considered given the importance of testing on resource utilization and the current understanding that lower socioeconomic status patients, many who may be uninsured, are disproportionately affected by COVID-19[35-37].

Unlike with testing, there is currently a lack of federal policies facilitating access to COVID-19 treatment via limiting cost-sharing. In the present study, we found that treatment contributed a larger portion of the reduction in mortality relative to diagnostic testing, highlighting the importance of ensuring access to treatments. Costs for current treatments are bundled as part of hospitalization costs and cost sharing will largely depend on insurance type. While many private payers have waived cost sharing for their members[38], some patients may still be vulnerable to high cost-sharing responsibilities such as those with high deductible insurance plans (that have not waived cost sharing) or the uninsured. For these patients who may delay seeking care due to cost, this may result in additional resource utilization due to missed opportunities to leverage future/existing treatments which may, for example, work in earlier stage disease and avoid the need for costlier ICU care and/or ventilator use. Furthermore, delays or avoiding care may also result in greater mortality[39, 40] and downstream productivity losses, accentuating the existing disparities in care. Future policies which ensure equitable access to hospital care and treatment should be considered.

Like all models, ours has limitations. First, the model assumes equal distribution of treatments and testing across the US. In reality, we know health care in the US is not always equally distributed relative to the number of cases in a region, so the impact of diagnostics and treatments may vary by individual health system. Thus, estimates of expected resource use relative to known availability of hospital/ICU beds and ventilators throughout the country should only be interpreted in aggregate across the country and is not necessarily reflective of the resource burden faced by individual health systems. While this model is based on historical data, for those wishing to estimate future local or regional scenarios for healthcare capacity planning, particularly as new advancements in the diagnosis and care of COVID-19 patients become available, we are happy to share the model, and it is available on request. Second, expanded testing scenarios assume that laboratory infrastructures are in place and the consumables required for testing are also widely available when in reality this may not be the case. Lastly, the model assumes public and private sector treatments and testing became available instantaneously on a single date when in actuality the availability of new diagnostics and treatments were spread out over time. For example, this may overestimate the impact of public sector contributions, as it is assumed both public sector testing capacity and treatments (dexamethasone) were available at scale since the beginning of the pandemic.

## Conclusion

Public and private sector efforts have provided substantial contributions to reducing COVID-19-related transmission, health care resource utilization, and mortality. Both diagnostic testing capacity and accuracy are important aspects to consider when identifying how to optimally deploy testing resources. Current COVID-19 treatment options are limited, yet provide significant contributions to reducing health care resource utilization and mortality through treatment benefits in the less severe hospitalized patients. Future combinations of treatments which impact different aspects of resource use may be optimal in reducing COVID-19-related pressure on healthcare system capacity. Policies that incentivize innovation and ensure equitable access to hospital care, testing, and treatment will be important to facilitate this.

## Data Availability

The datasets generated during and/or analyzed during the current study are available in the COVID Tracking Project repository, https://covidtracking.com/data/api. Programming code used to estimate model parameters is available on Github, https://github.com/Roche/covid-hcru-model.

https://github.com/Roche/covid-hcru-model

## Notes

### Competing Interest Statement

Daniel Sheinson, Caroline Solon, William Wong, and Anuj Shah are all salaried employees of Genentech; Mindy Cheng is a salaried employee of Roche Molecular Systems, Inc. David Elsea and Yang Meng received payment from Genentech for consulting fees. Daniel Sheinson, Caroline Solon, William Wong, and Mindy Cheng all hold Roche stock.

### Funding Statement

No external funding was received.

### Author Declarations

All datasets analyzed in this study are publicly available.

## References

1. John Hopkins University & Medicine. COVID-19 Dashboard. 2020 [cited 2020 10 Aug]; Available from: https://coronavirus.jhu.edu/map.html

2. Wu, Z. and J.M. McGoogan, Characteristics of and Important Lessons From the Coronavirus Disease 2019 (COVID-19) Outbreak in China: Summary of a Report of 72314 Cases From the Chinese Center for Disease Control and Prevention. JAMA, 2020. 323(13): p. 1239–1242.

3. Kim, L., et al., Risk Factors for Intensive Care Unit Admission and In-hospital Mortality among Hospitalized Adults Identified through the U.S. Coronavirus Disease 2019 (COVID-19)-Associated Hospitalization Surveillance Network (COVID-NET). Clin Infect Dis, 2020.

4. Adelman, D., Thousands Of Lives Could Be Saved In The US During The COVID-19 Pandemic If States Exchanged Ventilators. Health Aff (Millwood), 2020. 39(7): p. 1247–1252.

5. Emanuel, E.J., et al., Fair Allocation of Scarce Medical Resources in the Time of Covid-19. N Engl J Med, 2020. 382(21): p. 2049–2055.

6. Moghadas, S.M., et al., Projecting hospital utilization during the COVID-19 outbreaks in the United States. Proc Natl Acad Sci U S A, 2020. 117(16): p. 9122–9126.

7. Uppal, A., et al., Critical Care And Emergency Department Response At The Epicenter Of The COVID-19 Pandemic. Health Aff (Millwood), 2020. 39(8): p. 1443–1449.

8. Argenziano, M.G., et al., Characterization and clinical course of 1000 patients with coronavirus disease 2019 in New York: retrospective case series. BMJ, 2020. 369: p. m1996.

9. Cummings, M.J., et al., Epidemiology, clinical course, and outcomes of critically ill adults with COVID-19 in New York City: a prospective cohort study. Lancet, 2020. 395(10239): p. 1763–1770.

10. Seneff, M.G., et al., Predicting the duration of mechanical ventilation. The importance of disease and patient characteristics. Chest, 1996. 110(2): p. 469–79.

11. Aiken, L.H., et al., Hospital nurse staffing and patient mortality, nurse burnout, and job dissatisfaction. JAMA, 2002. 288(16): p. 1987–93.

12. Coster, S., What is the impact of professional nursing on patients’ outcomes globally? An overview of research evidence. Int J Nurs Stud, 2018. 78: p. 76–83.

13. Gupta, S., et al., Factors Associated With Death in Critically Ill Patients With Coronavirus Disease 2019 in the US. JAMA Intern Med, 2020.

14. Lee, A., et al., Are high nurse workload/staffing ratios associated with decreased survival in critically ill patients? A cohort study. Ann Intensive Care, 2017. 7(1): p. 46.

15. Weissman, G.E., et al., Locally Informed Simulation to Predict Hospital Capacity Needs During the COVID-19 Pandemic. Ann Intern Med, 2020. 173(1): p. 21–28.

16. National Institutes of Health. Accelerating COVID-19 Therapeutic Innovations and Vaccines (ACTIV). 2020 [cited 09 Sept 2020]; Available from: https://www.nih.gov/research-training/medical-research-initiatives/activ.

17. The COVID Tracking Project. Our Data. 2020 [cited 22 Aug 2020]; Available from: https://covidtracking.com/data.

18. Mizumoto, K., et al., Estimating the asymptomatic proportion of coronavirus disease 2019 (COVID-19) cases on board the Diamond Princess cruise ship, Yokohama, Japan, 2020. Euro Surveill, 2020. 25(10).

19. World Health Organization. Report of the WHO-China Joint Mission on coronavirus disease 2019 (COVID-19). 2020 [cited 04/20/2020]; Available from: https://www.who.int/publications/i/item/report-of-the-who-china-joint-mission-on-coronavirus-disease-2019-(covid-19).

20. Liu, J. and M. West, Combined Parameter and State Estimation in Simulation-Based Filtering, in Sequential Monte Carlo Methods in Practice, A. Doucet, N. de Freitas, and N. Gordon, Editors. 2001, Springer New York: New York, NY. p. 197–223.

21. R Core Team. R: A Language and Environment for Statistical Computing. 2019; Available from: https://www.R-project.org/.

22. Centers for Medicare & Medicaid Services. CMS-Ruling 2020-1-R. 2020 [cited 10 Aug 2020]; Available from: https://www.cms.gov/files/document/cms-2020-01-r.pdf.

23. Association for Molecular Pathology. SARS-CoV-2 Molecular Testing - Summary of Recent SARS-CoV-2 Molecular Testing Survey. 2020 [cited 14 Aug 2020]; Available from: https://www.amp.org/AMP/assets/AMP_SARS-CoV-2_Survey_Report_FINAL.pdf?pass=57.

24. Recovery Collaborative Group, et al., Dexamethasone in Hospitalized Patients with Covid-19 - Preliminary Report. N Engl J Med, 2020.

25. Beigel, J.H., et al., Remdesivir for the Treatment of Covid-19 - Preliminary Report. N Engl J Med, 2020.

26. Dyer, O., Covid-19: Remdesivir has little or no impact on survival, WHO trial shows. BMJ, 2020. 371: p. m4057.

27. Pan, H., et al., Repurposed antiviral drugs for COVID-19 –interim WHO SOLIDARITY trial results. medRxiv, 2020: p. 2020.10.15.20209817.

28. Healthcare Cost and Utilization Project. 2017 cost data for DRG 207 (Respiratory system diagnosis w ventilator support 96+ hours) and DRG 179 (Respiratory infections & inflammations w/o cc/mcc). [cited 28 Aug 2020]; Available from: https://hcupnet.ahrq.gov/#setup.

29. Centers for Medicare & Medicaid Services. Medicare Part D Drug Spending Dashboard & Data. 2020 [cited 14 Sept 2020]; Available from: https://www.cms.gov/Research-Statistics-Data-and-Systems/Statistics-Trends-and-Reports/Information-on-Prescription-Drugs/MedicarePartD.

30. Gilead Sciences. Press Release - An Open Letter from Daniel O-Day, Chairman & CEO, Gilead Sciences. 2020 [cited 29 Jun 2020]; Available from: https://www.gilead.com/news-and-press/press-room/press-releases/2020/6/an-open-letter-from-daniel-oday-chairman--ceo-gilead-sciences.

31. National Institutes of Health. Coronavirus Disease 2019 (COVID-19) Treatment Guidelines. 2020 [cited 09/21/2020]; Available from: https://www.covid19treatmentguidelines.nih.gov/.

32. American Clinical Laboratory Association. Press Release 07/14/2020 - ACLA Update on PCR Testing Capacity for Covid 19. 2020 [cited 14 Sept 2020]; Available from: https://www.acla.com/acla-update-on-pcr-testing-capacity-for-covid-19/.

33. Milken Institute. COVID-19 Treatment and Vaccine Tracker. 2020 [cited 28 Aug 2020]; Available from: https://covid-19tracker.milkeninstitute.org/.

34. Centers for Medicare & Medicaid Services. FAQs about Families First Coronavirus Response Act and Coronavirus Aid, Relief, and Economic Security implementation Part 43. 2020 [cited 26 Aug 2020]; Available from: https://www.cms.gov/files/document/FFCRA-Part-43-FAQs.pdf.

35. Centers for Disease Control and Prevention. Health equity considerations and racial and ethnic minority groups. 2020 [cited 28 Aug 2020]; Available from: https://www.cdc.gov/coronavirus/2019-ncov/community/health-equity/race-ethnicity.html.

36. Krouse, H.J., COVID-19 and the Widening Gap in Health Inequity. Otolaryngol Head Neck Surg, 2020. 163(1): p. 65–66.

37. Shadmi, E., et al., Health equity and COVID-19: global perspectives. Int J Equity Health, 2020. 19(1): p. 104.

38. Kudman, J.M. D; Kurani, N; et al,. Cost-sharing waivers and premium relief by private plans in response to COVID-19. 2020 [cited 28 Aug 2020]; Available from: https://www.healthsystemtracker.org/brief/cost-sharing-waivers-and-premium-relief-by-private-plans-in-response-to-covid-19/.

39. Giammaria, D.a.A.P., Can early treatment of patients with risk factors contribute to managing the COVID-19 pandemic? J Glob Health, 2020. 10(1): p. 010377.

40. Sun, Q., et al., Lower mortality of COVID-19 by early recognition and intervention: experience from Jiangsu Province. Ann Intensive Care, 2020. 10(1): p. 33.

