## Supplemental Appendix for "Impact of public and private sector COVID-19 diagnostics and treatments on US healthcare resource utilization"

#### 1 Model Description

Following Moghadas et al. (2020), we specify an age-stratified compartmental model of the COVID-19 pandemic in which we track the number of susceptible ($S_t^a$), exposed ($E_t^a$), mildly symptomatic ($A_t^{i,a}$), severely symptomatic ($I_t^{i,h,a}$), hospitalized ($H_t^{c,v,a}$), deceased ($D_t^a$), and recovered ($R_t^a$) individuals in age group $a$ at time $t$, where $i \in \{0,1\}$ indicates whether individuals are self-isolated, $h \in \{0,1\}$ indicates whether individuals require hospitalization, $c \in \{0,1\}$ indicates whether hospital patients get admitted to the intensive-care unit (ICU), and $v \in \{0,1\}$ indicates whether hospital patients require mechanical ventilation. We assumed that mechanical ventilation is only present in the ICU, so that $H_t^{0,1,a}$ does not exist.

Furthermore, we incorporate additional states that represent the number of patients that are infected but asymptomatic, $\tilde{A}_t^{i,a}$, and the number of patients under testing (from the time the patient goes to get tested to the time the test result is made available), $T_t^{s,h,a}$, where $s \in \{0,1,2\}$ is an indicator of whether the patient is asymptomatic (0), mildly symptomatic (1) or severely symptomatic (2) and $h \in \{0,1\}$ again indicates whether patients require hospitalization. We assumed that asymptomatic and mildly symptomatic patients will not require hospitalization (so that $T_t^{0,1,a}$ and $T_t^{1,1,a}$ do not exist). After adding these additional states, the original $E_t^a$ represent exposed patients who will become infectious, while the original infectious states ($\tilde{A}_t^{i,a}$, $A_t^{i,a}$, $I_t^{i,h,a}$) represent infected patients who are not under testing. The model structure is pictured in Figure S1.

We also included additional dynamic model states to represent changes in the effective transmission rate over time (e.g. due to social distancing), $\xi_t$, and changes in the proportion of mild and severe patients getting tested over time ($w_t^A$ and $w_t^I$ for mild and severe, respectively).

Mathematically, $S_t^a$, $E_t^a$, $\tilde{A}_t^{i,a}$, $A_t^{i,a}$, $I_t^{i,h,a}$, $T_t^{s,h,a}$, $H_t^{c,v,a}$, $D_t^a$, $R_t^a$, and $\xi_t$ are all nonnegative for all $t$, while $w_t^A$ and $w_t^I$ are between 0 and 1 for all $t$. When monitoring the pandemic, the true $S_t^a$, $E_t^a$, $\tilde{A}_t^{i,a}$, $A_t^{i,a}$, $I_t^{i,h,a}$, $T_t^{s,h,a}$, $H_t^{c,v,a}$, $D_t^a$, and $R_t^a$, $\xi_t$, $w_t^A$, and $w_t^I$ are unknown and regarded as hidden states of the model.

The model is also governed by fixed parameters related to disease transmission, behavior and symptoms, testing, and hospitalization. These are described in Table S1.

Lastly, the unknown states and fixed parameters were assumed to be related to observed data from The COVID Tracking Project (covidtracking.com). The data include the reported number of positive tests ($p_t$), patients currently hospitalized ($h_t$), patients in the ICU ($i_t$), patients on mechanical ventilation ($v_t$), and cumulative deaths ($d_t$) at time $t$. Given the states, fixed parameters, and observed data defined above, we construct a state-space model of the COVID-19 pandemic in the US, in which an observation equation specifies how the observed data depend on the hidden state of the pandemic and the state equation describes how the pandemic evolves over time.

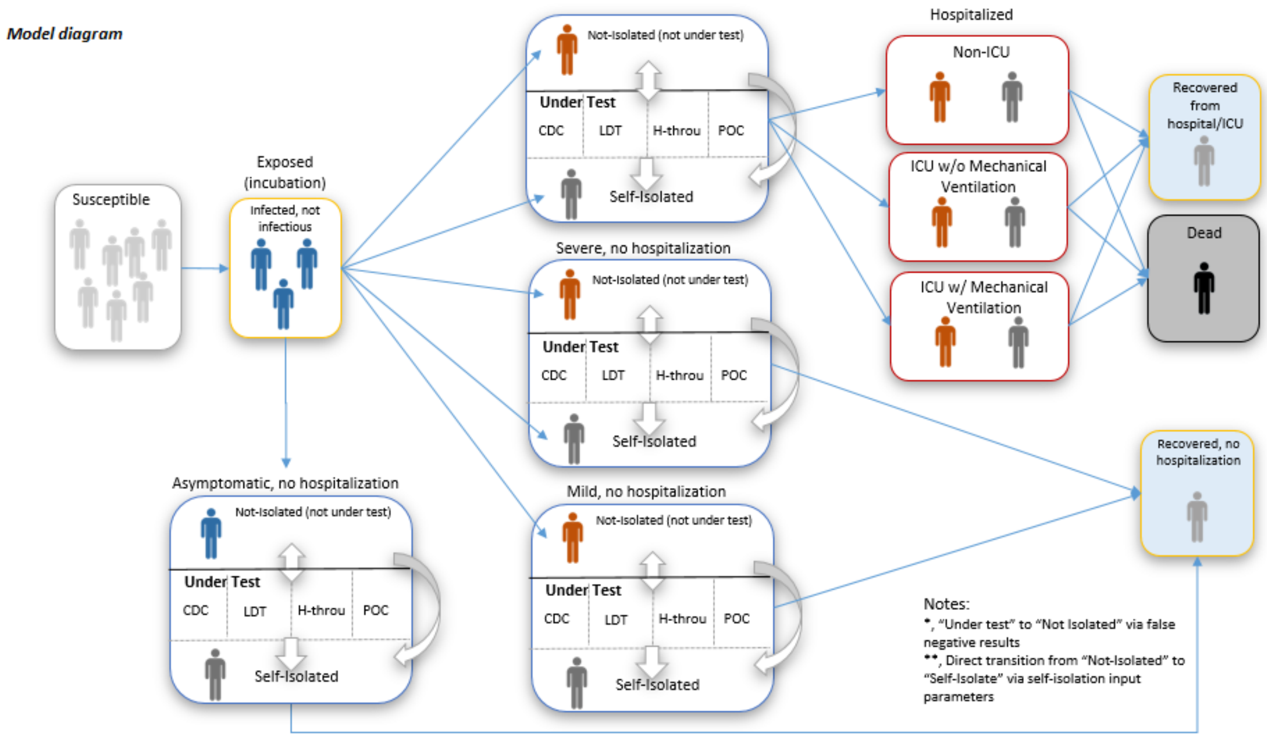


Figure S1: Compartmental model structure.

#### 1.1 State equation

First, we describe the state equation. Let

$$
\begin{aligned}
x_t^a = (&S_t^a, E_t^a,\\
&\tilde{A}_t^{0,a}, \tilde{A}_t^{1,a}, A_t^{0,a}, A_t^{1,a},\\
&I_t^{0,0,a}, I_t^{1,0,a}, I_t^{0,1,a}, I_t^{1,1,a},\\
&T_t^{0,0,a}, T_t^{1,0,a}, T_t^{2,0,a}, T_t^{2,1,a},\\
&H_t^{0,0,a}, H_t^{1,0,a}, H_t^{1,1,a},\\
&D_t^a, R_t^a,\\
&\xi_t, w_t^A, w_t^I)'
\end{aligned}
$$

be the state of the pandemic for age group $a$ at time $t$, and let $x_t = (x_t^{1\prime}, x_t^{2\prime}, \ldots, x_t^{\lambda\prime})'$, where $\lambda$ is the number of age groups.

For the testing turnaround rate (i.e. inverse of the test system delay) and testing sensitivity parameters $\varphi$ and $\omega$, we assumed these represent the weighted average of $L$ available tests on the market, such that

$$
\varphi = \sum_{j=1}^{L} s_j \varphi_j \qquad \omega = \sum_{j=1}^{L} s_j \omega_j
$$

where $\varphi_j$, $\omega_j$, and $s_j \in [0, 1]$ represent the turnaround rate, sensitivity, and market share, respectively, of test $j$, with $s_1 + s_2 + \cdots s_L = 1$.

Let the likelihood of coming in contact with an infectious individual in the community $(C_t^a)$ versus while in

| Parameter | Description |
|---|---|
| ***Transmission parameters*** | |
| $P_a \geq 0$ | population size for age group $a$ |
| $\beta \geq 0$ | transmission rate per contact |
| $M_{a,c} \geq 0$ | community contact rate per day between people in age groups $a$ and $c$ |
| $\tilde{M}_{a,c} \geq 0$ | household (isolation) contact rate per day between people in age groups $a$ and $c$ |
| $\kappa_M \in [0,1]$ | relative infectivity of mild versus severe illness |
| $\kappa_A \in [0,1]$ | relative infectivity of asymptomatic versus severe illness |
| $\nu \geq 0$ | symptom onset rate per day among infected (inverse of the incubation period) |
| ***Symptoms and behavioral parameters*** | |
| $\zeta_a \in [0,1]$ | proportion of infected individuals in age group $a$ that are asymptomatic |
| $\theta_a \in [0,1]$ | proportion of cases in age group $a$ exhibiting mild symptoms |
| $\gamma \geq 0$ | recovery rate per day from symptom onset |
| $q \in [0,1]$ | proportion of severe cases who practice self-isolation upon symptom onset |
| $f_I \in [0,1]$ | weight associated with self-isolation after symptom onset for severe cases |
| $f_A \in [0,1]$ | weight associated with self-isolation after symptom onset for mild cases |
| $f_T \in [0,1]$ | proportion of cases that self-isolate while waiting for test results |
| $\tau_I \geq 0$ | self-isolation rate per day for severe cases after symptom onset |
| $\tau_A \geq 0$ | self-isolation rate per day for mild cases after symptom onset |
| $\phi_\xi \in (0,1)$ | autocorrelation parameter for the change in transmission rate over time |
| $\sigma_\xi \geq 0$ | white noise standard deviation of the change in transmission rate over time |
| ***Testing parameters*** | |
| $w_s \in [0,1]$ | proportion of asymptomatic cases that get tested |
| $r_I \geq 0$ | rate per day of testing for severe cases |
| $r_A \geq 0$ | rate per day of testing for asymptomatic and mild cases |
| $\varphi \geq 0$ | daily turnaround rate of testing |
| $\omega \geq 0$ | sensitivity of testing |
| $\phi_I \in (0,1)$ | autocorrelation parameter for the change in testing of severe cases over time |
| $\phi_A \in (0,1)$ | autocorrelation parameter for the change in testing of mild cases over time |
| $\sigma_w \geq 0$ | white noise standard deviation of the change in testing of severe and mild cases over time |
| $\alpha \geq 0$ | upper limit of overall daily testing capacity |
| $p_\alpha \in [0,1]$ | proportion of testing capacity applied to the infected population |
| ***Hospitalization parameters*** | |
| $\delta \geq 0$ | hospitalization rate per day |
| $\rho_a \in [0,1]$ | proportion of severe cases in age group $a$ requiring hospitalization |
| $c_a \in [0,1]$ | proportion of hospitalized cases in age group $a$ admitted to the ICU |
| $v \in [0,1]$ | proportion of patients admitted to the ICU that require mechanical ventilation |
| $\mu_h \geq 0$ | rate per day of death for hospitalized patients not admitted to the ICU |
| $\mu_c \geq 0$ | rate per day of death for patients admitted to the ICU but not on mechanical ventilation |
| $\mu_v \geq 0$ | rate per day of death for patients on mechanical ventilation |
| $m_h \in [0,1]$ | weight associated with death for hospitalized patients not admitted to the ICU |
| $m_c \in [0,1]$ | weight associated with death for ICU patients not on mechanical ventilation |
| $m_v \in [0,1]$ | weight associated with death for patients on mechanical ventilation |
| $\psi_h \geq 0$ | rate per day of recovery for hospitalized patients not admitted to the ICU |
| $\psi_c \geq 0$ | rate per day of recovery for patients admitted to the ICU but not on mechanical ventilation |
| $\psi_v \geq 0$ | rate per day of recovery for patients on mechanical ventilation |

Table S1: Definitions of fixed parameters.

isolation ($O_t^a$) be defined by the following:

$$C_t^a = \frac{I_t^{0,0,a} + I_t^{0,1,a} + \kappa_M A_t^{0,a} + \kappa_A \tilde{A}_t^{0,a} + (1-f_T)\left(T_t^{2,0,a} + T_t^{2,1,a} + \kappa_M T_t^{1,0,a} + \kappa_A T_t^{0,0,a}\right)}{P_a}$$

$$O_t^a = \frac{I_t^{1,0,a} + I_t^{1,1,a} + \kappa_M A_t^{1,a} + \kappa_A \tilde{A}_t^{1,a} + f_T\left(T_t^{2,0,a} + T_t^{2,1,a} + \kappa_M T_t^{1,0,a} + \kappa_A T_t^{0,0,a}\right)}{P_a}$$

Let $T_t = \sum_{j=1}^{\lambda}\left(T_t^{0,0,j} + T_t^{1,0,j} + T_t^{2,0,j} + T_t^{2,1,j}\right)$ be the total number of infected individuals under testing at time $t$ and define

$$\tilde{T}_t^{0,0,a} = \min\left(T_t^{0,0,a}, p_\alpha \alpha(T_t^{0,0,a} \,/\, T_t)\right) \qquad \tilde{T}_t^{1,0,a} = \min\left(T_t^{1,0,a}, p_\alpha \alpha(T_t^{1,0,a} \,/\, T_t)\right)$$
$$\tilde{T}_t^{2,0,a} = \min\left(T_t^{2,0,a}, p_\alpha \alpha(T_t^{2,0,a} \,/\, T_t)\right) \qquad \tilde{T}_t^{2,1,a} = \min\left(T_t^{2,1,a}, p_\alpha \alpha(T_t^{2,1,a} \,/\, T_t)\right)$$

as the number of individuals in each testing state that are able to receive testing results on a given day. We describe the evolution of the disease states from time $t$ to $t+1$ deterministically by the following equations:

$$S_{t+1}^a = S_t^a - \xi_t \beta S_t^a \left(\sum_{j=1}^{\lambda} M_{a,j} C_t^a + \sum_{j=1}^{\lambda} \tilde{M}_{a,j} O_t^a\right) \tag{1}$$

$$E_{t+1}^a = E_t^a + \xi_t \beta S_t^a \left(\sum_{j=1}^{\lambda} M_{a,j} C_t^a + \sum_{j=1}^{\lambda} \tilde{M}_{a,j} O_t^a\right) - \nu E_t^a \tag{2}$$

$$\tilde{A}_{t+1}^{0,a} = \tilde{A}_t^{0,a} + \zeta_a \nu E_t^a + \phi(1-\omega)\tilde{T}_t^{0,0,a} - w_s r_A \tilde{A}_t^{0,a} - (1-w_s)\gamma \tilde{A}_t^{0,a} \tag{3}$$

$$\tilde{A}_{t+1}^{1,a} = \tilde{A}_t^{1,a} + \phi\omega \tilde{T}_t^{0,0,a} - \gamma \tilde{A}_t^{1,a} \tag{4}$$

$$\begin{aligned} A_{t+1}^{0,a} &= A_t^{0,a} + (1-\zeta_a)\theta_a \nu E_t^a + \phi(1-\omega)\tilde{T}_t^{1,0,a} \\ &\quad - w_t^A r_A A_t^{0,a} - (1-w_t^A) f_A \tau_A A_t^{0,a} - (1-w_t^A)(1-f_A)\gamma A_t^{0,a} \end{aligned} \tag{5}$$

$$A_{t+1}^{1,a} = A_t^{1,a} + f_A \tau_A A_t^{0,a} + \phi\omega \tilde{T}_t^{1,0,a} - \gamma A_t^{1,a} \tag{6}$$

$$\begin{aligned} I_{t+1}^{0,0,a} &= I_t^{0,0,a} + (1-\zeta_a)(1-\theta_a)(1-q)(1-\rho_a)\nu E_t^a + \phi(1-\omega)\tilde{T}_t^{2,0,a} \\ &\quad - w_t^I r_I I_t^{0,0,a} - (1-w_t^I) f_I \tau_I I_t^{0,0,a} - (1-w_t^I)(1-f_I)\gamma I_t^{0,0,a} \end{aligned} \tag{7}$$

$$I_{t+1}^{1,0,a} = I_t^{1,0,a} + (1-\zeta_a)(1-\theta_a) q (1-\rho_a)\nu E_t^a + (1-w_t^I) f_I \tau_I I_t^{0,0,a} + \phi\omega \tilde{T}_t^{2,0,a} - \gamma I_t^{1,0,a} \tag{8}$$

$$\begin{aligned} I_{t+1}^{0,1,a} &= I_t^{0,1,a} + (1-\zeta_a)(1-\theta_a)(1-q)\rho_a \nu E_t^a + \phi(1-\omega)\tilde{T}_t^{2,1,a} \\ &\quad - w_t^I r_I I_t^{0,1,a} - (1-w_t^I) f_I \tau_I I_t^{0,1,a} - (1-w_t^I)(1-f_I)\delta I_t^{0,1,a} \end{aligned} \tag{9}$$

$$I_{t+1}^{1,1,a} = I_t^{1,1,a} + (1-\zeta_a)(1-\theta_a) q \rho_a \nu E_t^a + (1-w_t^I) f_I \tau_I I_t^{0,1,a} + \phi\omega \tilde{T}_t^{2,1,a} - \delta I_t^{1,1,a} \tag{10}$$

$$T_{t+1}^{0,0,a} = T_t^{0,0,a} + w_s r_A \tilde{A}_t^{0,a} - \phi \tilde{T}_t^{0,0,a} - \gamma T_t^{0,0,a} \tag{11}$$

$$T_{t+1}^{1,0,a} = T_t^{1,0,a} + w_t^A r_A A_t^{0,a} - \phi \tilde{T}_t^{1,0,a} - \gamma T_t^{1,0,a} \tag{12}$$

$$T_{t+1}^{2,0,a} = T_t^{2,0,a} + w_t^I r_I I_t^{0,0,a} - \phi \tilde{T}_t^{2,0,a} - \gamma T_t^{2,0,a} \tag{13}$$

$$T_{t+1}^{2,1,a} = T_t^{2,1,a} + w_t^I r_I I_t^{0,1,a} - \phi \tilde{T}_t^{2,1,a} - \delta T_t^{2,1,a} \tag{14}$$

$$\begin{aligned} H_{t+1}^{0,0,a} &= H_t^{0,0,a} + (1-c_a)(1-w_t^I)(1-f_I)\delta I_t^{0,1,a} + (1-c_a)\delta I_t^{1,1,a} + (1-c_a)\delta T_t^{2,1,a} \\ &\quad - \left(m_h \mu_h + (1-m_h)\psi_h\right) H_t^{0,0,a} \end{aligned} \tag{15}$$

$$\begin{aligned} H_{t+1}^{1,0,a} &= H_t^{1,0,a} + (1-v)c_a(1-w_t^I)(1-f_I)\delta I_t^{0,1,a} + (1-v)c_a \delta I_t^{1,1,a} + (1-v)c_a \delta T_t^{2,1,a} \\ &\quad - \left(m_c \mu_c + (1-m_c)\psi_c\right) H_t^{1,0,a} \end{aligned} \tag{16}$$

$$H_{t+1}^{1,1,a} = H_t^{1,1,a} + v c_a (1-w_t^I)(1-f_I)\delta I_t^{0,1,a} + v c_a \delta I_t^{1,1,a} + v c_a \delta T_t^{1,1,a} - \left(m_v \mu_v + (1-m_v)\psi_v\right) H_t^{1,1,a} \tag{17}$$

$$D_{t+1}^a = D_t^a + m_h \mu_h H_t^{0,0,a} + m_c \mu_c H_t^{1,0,a} + m_v \mu_v H_t^{1,1,a} \tag{18}$$

$$\begin{aligned} R_{t+1}^a &= R_t^a + (1-w_t^A)(1-f_A)\gamma A_t^{0,a} + \gamma \tilde{A}_t^{1,a} + (1-w_t^A)(1-f_A)\gamma A_t^{0,a} \\ &\quad + \gamma A_t^{1,a} + (1-w_t^I)(1-f_I)\gamma I_t^{0,0,a} + \gamma I_t^{1,0,a} + \gamma T_t^{0,0,a} + \gamma T_t^{1,0,a} + \gamma T_t^{2,0,a} \\ &\quad + (1-m_h)\psi_h H_t^{0,0,a} + (1-m_c)\psi_c H_t^{1,0,a} + (1-m_v)\psi_v H_t^{1,1,a} \end{aligned} \tag{19}$$

The evolution of the dynamic states $\xi_t$, $w_t^A$, and $w_t^I$ from time $t$ to $t+1$ follow the constrained first-order autoregressive (AR1) processes

$$\xi_{t+1} = \max(\phi_\xi \xi_t + \epsilon_\xi, 0) \tag{20}$$

$$w_{t+1}^A = \min\left(\max(\phi_A(w_t^A - 1) + \epsilon_w + 1, 0), 1\right) \tag{21}$$

$$w_{t+1}^I = \min\left(\max(\phi_I(w_t^I - 1) + \epsilon_w + 1, 0), 1\right) \tag{22}$$

where $\epsilon_\xi$ and $\epsilon_w$ are independent, normally distributed random errors with mean 0 and standard deviations $\sigma_\xi$ and $\sigma_w$, respectively. The AR1 processes are defined such that they are stationary and constrained to be either nonnegative ($\xi_t$) or bounded by $(0,1)$ ($w_t^A$ and $w_t^I$). The asymptotic means $\mathrm{E}(\xi_t) = 0$ and $\mathrm{E}(w_t^A) = \mathrm{E}(w_t^I) = 1$ are such that the effective transmission rate approaches 0 and the proportion of mild and severe cases tested approaches 100% in the long run.

##### 1.2 Observation equation

Let $y_t = (p_t, h_t, i_t, v_t, d_t)'$ be the observed data from The COVID Tracking Project, and let

$$T_t^p = \phi\omega \left[ \sum_{j=1}^{\lambda} \left( \tilde{T}_t^{0,0,j} + \tilde{T}_t^{1,0,j} + \tilde{T}_t^{2,0,j} + \tilde{T}_t^{2,1,j} \right) \right]$$

be the number of new positive tests at time $t$. We model the log of the observations by

$$\log p_t = \log T_t^p + \eta_p \tag{23}$$

$$\log h_t = \log \left[ \sum_{j=1}^{\lambda} \left( H_t^{0,0,j} + H_t^{1,0,j} + H_t^{1,1,j} \right) \right] + \eta_h \tag{24}$$

$$\log i_t = \log \left[ \sum_{j=1}^{\lambda} \left( H_t^{1,0,j} + H_t^{1,1,j} \right) \right] + \eta_i \tag{25}$$

$$\log v_t = \log \left[ \sum_{j=1}^{\lambda} H_t^{1,1,j} \right] + \eta_v \tag{26}$$

$$\log d_t = \log \left[ \sum_{j=1}^{\lambda} D_t^j \right] + \eta_d \tag{27}$$

where $\eta_p$, $\eta_h$ , $\eta_i$ , $\eta_v$, and $\eta_d$ are independent, normally distributed random errors with mean 0 and standard deviations $\sigma_p$, $\sigma_h$, $\sigma_i$, $\sigma_v$, and $\sigma_d$, respectively. The data are modelled on the log scale in order to constrain stochastic observations to be positive.

#### 2 Model Calibration

Having formulated the state-space model, let $\theta$ represent all unknown fixed parameters. Then, we specify the likelihood of an observation $y_t$ given the current state $x_t$ and fixed parameters $\theta$ by $p(y_t|x_t, \theta)$, which according to equations 23 through 27 follows a log-normal density with mean $F_t$ and covariance matrix $\Sigma_t$ defined on the log-scale by

$$F_t = \begin{pmatrix} \log T_t^p \\ \log \left[ \sum_{j=1}^{\lambda} \left( H_t^{0,0,j} + H_t^{1,0,j} + H_t^{1,1,j} \right) \right] \\ \log \left[ \sum_{j=1}^{\lambda} \left( H_t^{1,0,j} + H_t^{1,1,j} \right) \right] \\ \log \left[ \sum_{j=1}^{\lambda} H_t^{1,1,j} \right] \\ \log \left[ \sum_{j=1}^{\lambda} D_t^j \right] \end{pmatrix} \qquad \Sigma_t = \begin{pmatrix} \sigma_p^2 & 0 & 0 & 0 & 0 \\ 0 & \sigma_h^2 & 0 & 0 & 0 \\ 0 & 0 & \sigma_i^2 & 0 & 0 \\ 0 & 0 & 0 & \sigma_v^2 & 0 \\ 0 & 0 & 0 & 0 & \sigma_d^2 \end{pmatrix}$$

The probability density of the future state $x_{t+1}$ given the current state $x_t$ and fixed parameters $\theta$, denoted by $p(x_{t+1}|x_t, \theta)$, can be sampled from according to the deterministic state equations 1 through 19 and constrained AR1 processes given by equations 20 through 22.

Let $x_{1:t} = (x_1, \ldots, x_t)$ and $y_{1:t} = (y_1, \ldots, y_t)$. We calibrate the model by sequentially estimating the filtered distribution at time $t+1$, $p(x_{t+1}, \theta|y_{1:t+1})$, given an estimate of the filtered distribution at time $t$, $p(x_t, \theta|y_{1:t})$, and a new data point, $y_{t+1}$. Since the state equations are nonlinear functions of $x_t$ and $\theta$, a closed-form solution to $p(x_{t+1}, \theta|y_{1:t+1})$ cannot be obtained. Thus, we use the kernel density particle filter (KDPF) described by Liu and West (2001) to approximate $p(x_t, \theta|y_{1:t})$ for all $t$.

#### 2.1 Kernel density particle filter

The KDPF approximates $p(x_t, \theta|y_{1:t})$ via a weighted sample of $J$ particles, i.e.

$$p(x_t, \theta|y_{1:t}) \approx \sum_{j=1}^{J} w_t^{(j)} I\left(x_t^{(j)}, \theta^{(j)}\right) \tag{28}$$

where $\left(x_t^{(j)}, \theta^{(j)}\right)$ is the location of the $j^{\text{th}}$ particle at time $t$, $w_t^{(j)}$ is the weight of that particle with $\sum_{j=1}^{J} w_t^{(j)} = 1$, and $I(x, \theta)$ is the Dirac delta function that puts probability mass at $(x, \theta)$. To make the notation transparent, we introduce subscripts for our fixed parameters, e.g. $\theta_t^{(j)}$ represents the value for $\theta$ at time $t$ for particle $j$. This does not imply that the true $\theta$ is dynamic, but rather that particle $j$ can have different values for $\theta$ throughout time.

One advantage of the KDPF is that it attempts to avoid degeneration in particle values (i.e. all particles end up having the same value) for fixed parameters that can result from repeated sampling. To do so, a kernel density approximation to the distribution of fixed parameters is constructed so that fixed parameter values can be regenerated at each time $t$. Let $\bar{\theta}_t$ and $V_t$ be the weighted sample mean and weighted sample covariance matrix of $\theta_t^{(1)}, \ldots, \theta_t^{(J)}$. The KDPF uses a tuning parameter $\Delta$, a discount factor that takes values in $(0, 1)$, and two derived quantities $h^2 = 1 - ((3\Delta - 1)/2\Delta)^2$ and $a^2 = 1 - h^2$ that determine how smooth the kernel density approximation is.

Given an approximation to the filtered distribution at time $t$ as in equation 28, the KDPF provides an approximation to $p(x_{t+1}, \theta|y_{1:t+1})$ by the following steps:

1. For each particle $j$, set $m_t^{(j)} = a\theta_t^{(j)} + (1 - a)\bar{\theta}_t$ and calculate a point estimate of $x_{t+1}^{(j)}$ called $\mu_{t+1}^{(j)}$, e.g. $\mu_{t+1}^{(j)} = E\left(x_{t+1} \middle| x_t^{(j)}, \theta_t^{(j)}\right)$.

2. Calculate auxiliary weights and renormalize:

$$\tilde{g}_{t+1}^{(j)} = w_t^{(j)} p\left(y_{t+1} \middle| \mu_{t+1}^{(j)}, m_t^{(j)}\right) \qquad g_{t+1}^{(j)} = \tilde{g}_{t+1}^{(j)} \Big/ \sum_{l=1}^{J} \tilde{g}_{t+1}^{(l)}.$$

3. For each particle $j = 1, \ldots, J$,

   (a) Resample: sample an index $k \in \{1, \ldots, j, \ldots, J\}$ with associated probabilities $\left\{g_{t+1}^{(1)}, \ldots, g_{t+1}^{(j)}, \ldots, g_{t+1}^{(J)}\right\}$,

   (b) Regenerate the fixed parameters: sample $\theta_{t+1}^{(j)} \sim \mathrm{N}\left(m_t^{(k)}, h^2 V_t\right)$,

   (c) Propagate: sample $x_{t+1}^{(j)} \sim p\left(x_{t+1} \middle| x_t^{(k)}, \theta_{t+1}^{(j)}\right)$, and

   (d) Calculate weights and renormalize:

$$\tilde{w}_{t+1}^{(j)} = \frac{p\left(y_{t+1} \middle| x_{t+1}^{(j)}, \theta_{t+1}^{(j)}\right)}{p\left(y_{t+1} \middle| \mu_{t+1}^{(k)}, m_t^{(k)}\right)} \qquad w_{t+1}^{(j)} = \tilde{w}_{t+1}^{(j)} \Big/ \sum_{l=1}^{J} \tilde{w}_{t+1}^{(l)}.$$

In the above implementation of the KDPF, we use a normal kernel, where $\mathrm{N}(\mu, \Sigma)$ represents the normal distribution with mean $\mu$ and covariance matrix $\Sigma$. For the point estimate of the future state for particle $j$, $\mu_{t+1}^{(j)}$, we use the deterministic state equations 1 through 19 and the conditional expectation of the AR1 processes from equations 20 through 22:

$$\begin{aligned}
E\left(\tilde{\xi}_{t+1} \,|x_t, \theta_t\right) &= \max(\phi_\xi \tilde{\xi}_t, 0) \\
E\left(\tilde{w}_{t+1}^A \,|x_t, \theta_t\right) &= \min\left(\max(\phi_A(\tilde{w}_t^A - 1) + 1, 0), 1\right) \\
E\left(\tilde{w}_{t+1}^I \,|x_t, \theta_t\right) &= \min\left(\max(\phi_I(\tilde{w}_t^I - 1) + 1, 0), 1\right)
\end{aligned}$$

To use the KDPF with normal kernels, it is necessary to parameterize the fixed parameters so that their support is on the real line. We used the logarithm transformation for parameters that have positive support and the logit transformation for parameters in the interval (0,1).

#### 2.2 KDPF Implementation

The KDPF was run on data collected daily by The COVID Tracking Project for the whole US. The time period of data collection that was analyzed was from 1/22/2020 through 8/21/2020, and time iterations of the KDPF corresponded to days (such that $t = 0$ corresponded to 1/21/2020, $t = 1$ to 1/22/2020, and so on). Four age categories were used in the age-stratified compartmental model (0-19, 20-49, 50-64, 65 and over), with the population size of each age group taken from US census data (https://www2.census.gov/programs-surveys/popest/) and shown in Table S2.

In order to avoid particle degeneracy in the KDPF, $J = 500000$ particles were used. Following recommendations from Liu and West (2001) and Sheinson et al. (2014), the discount factor $\Delta$ was set to 0.99 and stratified resampling with an effective sample size threshold of 0.8 was implemented using R package `smcUtils` (Niemi, 2012). All hospitalization parameters shown in Table S1, the proportion of testing capacity applied to infected population ($p_\alpha$), and the observation standard deviation parameters $\sigma_p$, $\sigma_h$, $\sigma_i$, $\sigma_v$, and $\sigma_d$ were assumed to be unknown. All other fixed parameters were assumed to be known and set at fixed values shown in Table S2. The KDPF runs were initialized by assuming 1 exposed individual in each age group. The unknown fixed parameters and initial states of the AR1 processes were sampled from independent prior distributions shown in Table S4. The KDPF was implemented in R version 3.5.3 (R Core Team, 2019) and the code is available on Github (https://github.com/Roche/covid-hcru-model).

### 3 Results

Estimates of the marginal filtered distributions of unknown fixed parameters were summarized in terms of their posterior means and 95% credible intervals, calculated via monte carlo estimates from the weighted particle samples (Table S5). In addition, these were calculated for aggregates of model states (e.g. for combining age groups) by summing the values of individual states together for each particle. Figure S2 displays these estimates for resource utilization over time.

Particle trajectories (or traces) were calculated for each of the unknown states by tracing the most recent value of each particle back to each parent particle from which it was resampled. From these traces, a mean particle trace could be calculated by taking the average particle trace at each time point. Alternatively, the particle trace that minimizes the sum of squared distances (SSd) between observed and traced resource utilization could be determined. The SSd-minimizing particle trace combined with the assumed values of known fixed parameters and posterior mean estimates of the unknown fixed parameters were used to simulate a pandemic similar to the one observed in the US and to generate the various testing and treatment scenarios discussed in the main manuscript. Figure S3 displays the simulated trajectories for each of the resource utilization states presented in Figure S2. Figure S4 shows the mean, SSd-minimizing, and individual particle traces for changes in the time-varying effective transmission rate, proportion of mild cases tested over time, and proportion of severe cases tested over time. Table S6 shows assumptions around treatment efficacy and expanded molecular diagnostic testing and how they were used to modify fixed parameters to represent different model scenarios.

| Parameter | Value(s) | | | | Source |
|---|---|---|---|---|---|
| | Age 0-19 | Age 20-44 | Age 50-64 | Age 65+ | |
| ***Transmission parameters*** | | | | | |
| $P_a$ | 82500000 | 132000000 | 62700000 | 52800000 | US Census |
| $\beta$ | 0.0493 | | | | Moghadas et al. (2020) |
| $M_{0-19,a}$ | 9.76 | 3.77 | 1.51 | 0.60 | Moghadas et al. (2020) |
| $M_{20-49,a}$ | 3.77 | 9.43 | 3.05 | 0.70 | Moghadas et al. (2020) |
| $M_{50-64,a}$ | 1.51 | 3.05 | 2.96 | 0.76 | Moghadas et al. (2020) |
| $M_{65+,a}$ | 0.60 | 0.70 | 0.76 | 0.60 | Moghadas et al. (2020) |
| $\tilde{M}_{0-19,a}$ | 2.04 | 1.56 | 0.50 | 0.38 | Moghadas et al. (2020) |
| $\tilde{M}_{20-49,a}$ | 1.56 | 1.51 | 0.45 | 0.24 | Moghadas et al. (2020) |
| $\tilde{M}_{50-64,a}$ | 0.50 | 0.45 | 1.04 | 0.19 | Moghadas et al. (2020) |
| $\tilde{M}_{65+,a}$ | 0.38 | 0.24 | 0.19 | 0.64 | Moghadas et al. (2020) |
| $\kappa_M$ | 0.5 | | | | Moghadas et al. (2020) |
| $\kappa_A$ | 0.5 | | | | Assumption |
| $\nu \geq 0$ | 1 / 5.2 | | | | Moghadas et al. (2020) |
| ***Symptoms and behavioral parameters*** | | | | | |
| $\zeta_a$ | 0.179 | | | | Mizumoto et al. (2020) |
| $\theta_a$ | 0.973 | 0.900 | 0.648 | 0.466 | WHO (2020), scaled to Moghadas et al. (2020) |
| $\gamma$ | 1 / 4.6 | | | | Moghadas et al. (2020) |
| $q$ | 0.05 | | | | Moghadas et al. (2020) |
| $f_I$ | 0.8 | | | | Moghadas et al. (2020) |
| $f_A$ | 0.05 | | | | Moghadas et al. (2020) |
| $f_T$ | 0.7 | | | | Assumption |
| $\tau_I$ | 1 / 1 | | | | Moghadas et al. (2020) |
| $\tau_A$ | 1 / 2 | | | | Moghadas et al. (2020) |
| $\phi_\xi$ | 0.95 | | | | Assumption |
| $\sigma_\xi$ | 0.25 | | | | Assumption |
| ***Testing parameters*** | | | | | |
| $w_s$ | 0.1 | | | | Assumption |
| $r_I$ | 1 / 2 | | | | Assumption |
| $r_A$ | 1 / 3 | | | | Assumption |
| $\phi$ | 1 / 2.3 | | | | Average of multiple tests, see Table S3 |
| $\omega$ | 0.972 | | | | Average of multiple tests, see Table S3 |
| $\phi_I$ | 0.995 | | | | Assumption |
| $\phi_A$ | 0.99 | | | | Assumption |
| $\sigma_w$ | 0.1 | | | | Assumption |
| $\alpha$ | 1122898 | | | | Market research, US Census data |

Table S2: Assumed values of fixed parameters.

| Test system | Market share ($s_j$) | Daily Throughput (range) | Sensitivity ($\omega_j$) | Turnaround time ($1/\phi_j$) | Source |
|---|---|---|---|---|---|
| HT | 43.2% | 470 - 2880 | 98.4% | 2 | Poljak et al. (2020)<br>Degli-Angeli et al. (2020)<br>Hogan et al. (2020) |
| POC | 11.8% | 32 | 93.0% | 1 | Zhen et al. (2020) |
| LDT | 25.0% | 16 | 97.2% | 3 | Set to average of commercial tests |
| CDC | 10.0% | 16 | 97.2% | 3 | Set to average of commercial tests |
| Other | 10.0% | 35 | 97.2% | 2.3 | Set to overall average |

Table S3: Testing capacity and system performance.

| Parameter | Distribution | Mean | Standard Deviation (hyperparameters) | Source for mean |
|---|---|---|---|---|
| $p_\alpha$ | Beta | 0.1 | 0.0420 ($shape = 5$, $rate = 45$) | Assumption |
| $\sigma_p$ | Gamma | 0.5 | 0.1000 ($shape = 25$, $rate = 50$) | Assumption |
| $\rho_{0-19}$ | Beta | 0.592 | 0.0688 ($a = 29.6$, $b = 20.4$) | Petrilli et al. (2020) |
| $\rho_{20-49}$ | Beta | 0.350 | 0.0668 ($a = 17.5$, $b = 32.5$) | Petrilli et al. (2020) |
| $\rho_{50-64}$ | Beta | 0.573 | 0.0693 ($a = 28.65$, $b = 21.35$) | Petrilli et al. (2020) |
| $\rho_{65+}$ | Beta | 0.698 | 0.0643 ($a = 34.9$, $b = 15.1$) | Petrilli et al. (2020) |
| $\sigma_h$ | Gamma | 0.5 | 0.1000 ($shape = 25$, $rate = 50$) | Assumption |
| $\delta$ | Gamma | 1 / 7 | 0.0535 ($shape = 7.14$, $rate = 50$) | Garg (2020) |
| $\psi_h$ | Gamma | 1 / 8.3 | 0.0491 ($shape = 6.02$, $rate = 50$) | Beigel et al. (2020) |
| $\psi_c$ | Gamma | 1 / 22 | 0.0302 ($shape = 2.27$, $rate = 50$) | Beigel et al. (2020) |
| $\psi_v$ | Gamma | 1 / 28 | 0.0267 ($shape = 1.79$, $rate = 50$) | Beigel et al. (2020) |
| $c_{0-19}$ | Beta | 0.152 | 0.1082 ($a = 1.52$, $b = 8.48$) | Richardson et al. (2020) |
| $c_{20-49}$ | Beta | 0.215 | 0.1239 ($a = 2.15$, $b = 7.85$) | Richardson et al. (2020) |
| $c_{50-64}$ | Beta | 0.215 | 0.1239 ($a = 2.15$, $b = 7.85$) | Richardson et al. (2020) |
| $c_{65+}$ | Beta | 0.237 | 0.1282 ($a = 2.37$, $b = 7.63$) | Richardson et al. (2020) |
| $\sigma_i$ | Gamma | 0.5 | 0.1000 ($shape = 25$, $rate = 50$) | Assumption |
| $v$ | Beta | 0.6 | 0.1477 ($a = 6$, $b = 4$) | Assumption |
| $\sigma_v$ | Gamma | 0.5 | 0.1000 ($shape = 25$, $rate = 50$) | Assumption |
| $m_h$ | Beta | 0.067 | 0.0754 ($a = 0.67$, $b = 9.33$) | Derived from Petrilli et al. (2020) |
| $m_c$ | Beta | 0.048 | 0.0645 ($a = 0.48$, $b = 9.52$) | Derived from Petrilli et al. (2020) |
| $m_v$ | Beta | 0.099 | 0.0900 ($a = 0.99$, $b = 9.01$) | Derived from Petrilli et al. (2020) |
| $\mu_h$ | Gamma | 1 / 9.7 | 0.0454 ($shape = 5.15$, $rate = 50$) | Moghadas et al. (2020) |
| $\mu_c$ | Gamma | 1 / 6.0 | 0.0577 ($shape = 8.33$, $rate = 50$) | ICNARC (2020) |
| $\mu_v$ | Gamma | 1 / 5.4 | 0.0609 ($shape = 9.26$, $rate = 50$) | Argenziano et al. (2020) |
| $\sigma_d$ | Gamma | 0.5 | 0.1000 ($shape = 25$, $rate = 50$) | Assumption |
| Initial state | Distribution | Mean | Standard Deviation (hyperparameters) | Source for mean |
| $\xi_0$ | Log-normal | 2 (log-scale) | 0.3 (log-scale) | Assumption |
| $w_0^A$ | Beta | 0.1 | 0.0299 ($a = 10$, $b = 90$) | Assumption |
| $w_0^I$ | Beta | 0.1 | 0.0299 ($a = 10$, $b = 90$) | Assumption |

Table S4: Prior distributions of unknown parameters.

| Parameter | Posterior Mean | | | |
|---|---|---|---|---|
| | Age 0-19 | Age 20-44 | Age 50-64 | Age 65+ |
| $p_\alpha$ | 0.0983 | | | |
| $\rho_a$ | 0.5961 | 0.4301 | 0.5228 | 0.7659 |
| $\delta$ | 0.1303 | | | |
| $\psi_h$ | 0.0580 | | | |
| $\psi_c$ | 0.0556 | | | |
| $\psi_v$ | 0.0484 | | | |
| $c_a$ | 0.1034 | 0.2058 | 0.1083 | 0.1420 |
| $v$ | 0.3556 | | | |
| $m_h$ | 0.2591 | | | |
| $m_c$ | 0.0887 | | | |
| $m_v$ | 0.0002 | | | |
| $\mu_h$ | 0.1441 | | | |
| $\mu_c$ | 0.1464 | | | |
| $\mu_v$ | 0.1249 | | | |

Table S5: Posterior means of unknown fixed parameters.

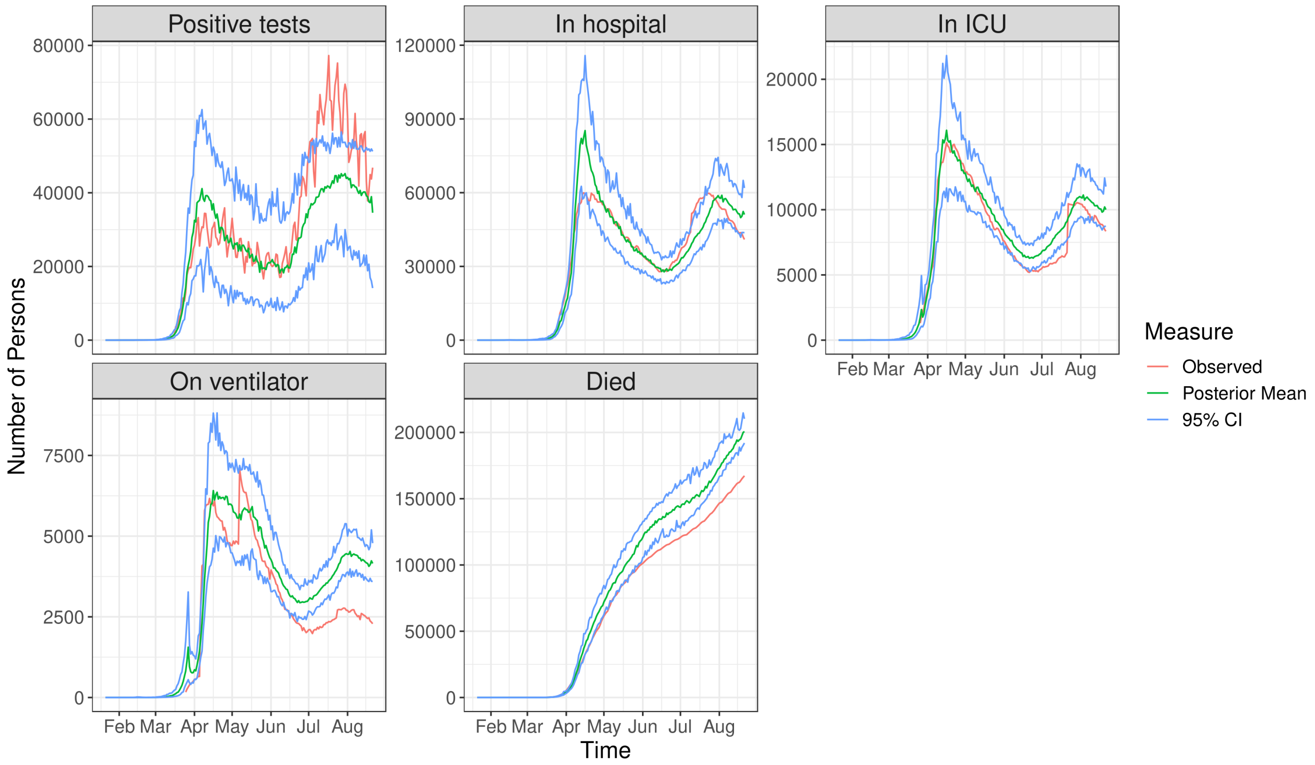


Figure S2: Posterior means (green lines) and 95% credible intervals (blue lines) of the marginal filtered distributions for resource utilization (panels) in the United States, alongside observed data (red lines).

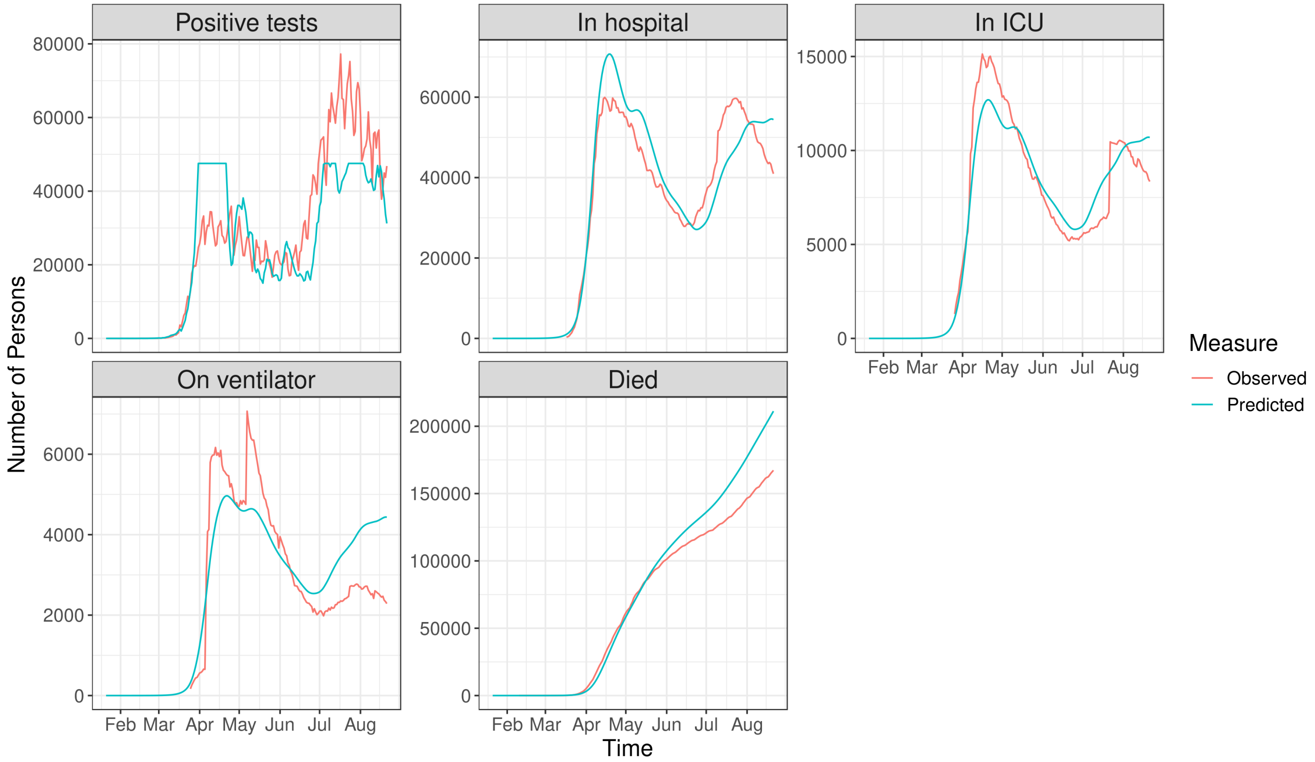


Figure S3: Simulated (blue lines) and observed (red lines) resource utilization (panels) in the United States.

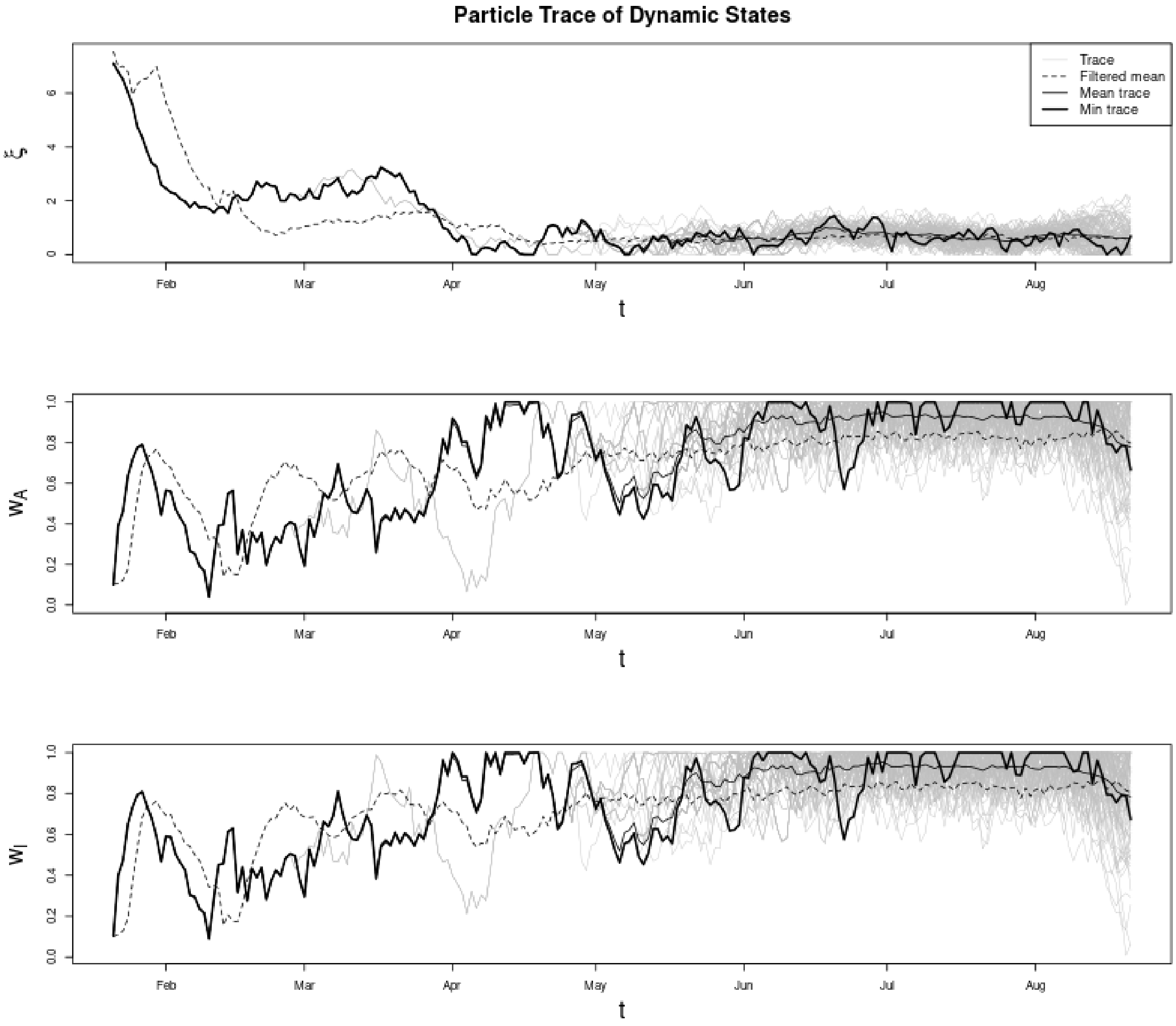


Figure S4: SSd-minimizing (thick black lines) and mean (thin black lines) particle traces of multiplicative changes in the effective transmission rate (top), proportion of mild cases tested (middle), and proportion of severe cases tested (bottom) over time in the United States. Dashed black lines denote the posterior means of the marginal filtered distributions and gray lines show the individual particle traces.

| Scenario | Modifications to model parameters (modified values denoted by *) |
|---|---|
| No testing capacity or treatment (Reference scenario) | -$\alpha^*$ set to 0 (No testing capacity)<br>-$w_s^*$ set to 0 (No asymptomatic testing)<br>-Mortality hazard ratios set to 1.25, 1.25, and 1.54 for non-ICU, ICU w/o vent, and ICU w/ vent patients, respectively (reverse the mortality benefit of dexamethasone, assuming 50% of patients treated since Day 1 (1/22/2020) of the model)<br>-Refer to Section 4 for detailed calculation on mortality |
| No private sector treatment or testing (Public sector only) | -$\alpha^*$ set to $s_j \times \alpha$, where $s_j$ set to 0 for all except LDT and CDC<br>-$s_j^*$'s renormalized to sum to 1 |
| Private sector treatment but not testing (Public + private sector) | -$\alpha^*$ set to $s_j \times \alpha$, where $s_j$ set to 0 for all except LDT and CDC<br>-$s_j^*$'s renormalized to sum to 1<br>-Length of stay for non-ICU patients reduced by 2.89 days, assuming 50% of patients are treated starting 6/1/2020<br>$\left(\psi_h^* = \frac{1}{\frac{1}{\psi_h} - 0.5 \times 2.89}\right)$<br>-Mortality hazard ratio for non-ICU patients of 0.28<br>-Refer to Section 4 for detailed calculation on mortality |
| Private sector testing but not treatment (Public + private sector) | -No change to model parameters |
| Private sector treatment and testing (Public + private sector) | -No change in testing parameters<br>-Length of stay for non-ICU patients reduced by 2.89 days, assuming 50% of patients are treated starting 6/1/2020<br>$\left(\psi_h^* = \frac{1}{\frac{1}{\psi_h} - 0.5 \times 2.89}\right)$<br>-Mortality hazard ratio for non-ICU patients of 0.28<br>-Refer to Section 4 for detailed calculation on mortality |

Table S6: Model scenario assumptions and corresponding modifications to model parameters.

### 4 Mortality Calculations

To apply the hazard ratio for the treatment effect, first the probability of death in the hospital ($p_m$) is calculated using the weights associated with mortality ($m$), mortality rates ($\mu$), and recovery rates ($\psi$). As in Moghadas et al. (2020), we calculate this probability as

$$p_m = \frac{m\mu}{m\mu + (1-m)\psi}$$

Then, we apply the hazard ratio for the mortality benefit to calculate the probability of death in the hospital among treated patients ($p_m^*$) according to

$$p_m^* = p_m - p_{trt}\left(p_m - \left[1 - (1-p_m)^{HR}\right]\right)$$

where $p_{trt}$ is the proportion of patients treated and $HR$ is the hazard ratio. For example, in our model scenarios incorporating private sector treatment benefit, $p_{trt}$ is assumed to be 0.5 and $HR$ is assumed to be 0.28 (Beigel et al., 2020). Finally, modified model weights ($m^*$) are then back-calculated according to

$$m^* = \frac{p_m^*\psi}{p_m^*\psi + (1-p_m^*)\mu}$$

### 5 Limitations

A few technical limitations regarding the model and estimation procedures are worth noting. First, observed data from The COVID Tracking Project are based on reported numbers from state and territory public health authorities. Not all states report each type of data, and they may vary in terms of their completeness. In addition, the reporting of the data is expected to lag behind actual cases. For more information on the data quality, refer to The COVID Tracking Project website (https://covidtracking.com/about-data).

In addition, the model may suffer from identifiability issues due to the large number of unknown states and fixed parameters. Thus, while calibration to the observed data may result in an accurate depiction of estimated resource use, testing, and mortality, individual model state and parameter estimates should be interpreted with caution, since multiple combinations of state and parameter values could result in similar estimates of resource use, testing, or mortality.

Lastly, simulated model scenarios assume that modifications to fixed parameters reflecting the impact of treatment and testing happen instantaneously at a single point in time, whereas a model that would allow for these parameters to change gradually over time is probably more reflective of reality. We made simplifying assumptions that 1. removal of public sector treatment and testing means that 50% of patients lose the clinical benefits of dexamethasone, and that LDT and CDC testing capacity are reduced to 0 starting at Day 1 of the model; and 2. addition of private sector treatment and testing means 50% of patients receive the clinical benefits of remdesivir, and that HT, POC, and other commercial testing capacity become available starting at 6/1/2020 (Day 132) of the model.

#### References


S. M. Moghadas, A. Shoukat, M. C. Fitzpatrick, C. R. Wells, P. Sah, A. Pandey, J. D. Sachs, Z. Wang, L. A. Meyers, B. H. Singer, A. P. Galvani, Projecting hospital utilization during the COVID-19 outbreaks in the United States, Proceedings of the National Academy of Sciences 117 (2020) 9122–9126.

J. Liu, M. West, Combined parameter and state estimation in simulation-based filtering, in: A. Doucet, J. F. G. De Freitas, N. J. Gordon (Eds.), Sequential Monte Carlo Methods in Practice, Springer-Verlag, New York, 2001, pp. 197–217.

D. M. Sheinson, J. Niemi, W. Meiring, Comparison of the performance of particle filter algorithms applied to tracking of a disease epidemic, Mathematical Biosciences 255 (2014) 21–32.

A. J. Niemi, smcUtils: Utility functions for sequential Monte Carlo, 2012. URL: `https://github.com/jarad/smcUtils`, R package version 0.2.2.

R Core Team, R: A Language and Environment for Statistical Computing, R Foundation for Statistical Computing, Vienna, Austria, 2019. URL: `https://www.R-project.org/`.

K. Mizumoto, K. Kagaya, A. Zarebski, G. Chowell, Estimating the asymptomatic proportion of coronavirus disease 2019 (COVID-19) cases on board the Diamond Princess cruise ship, Yokohama, Japan, 2020, Eurosurveillance 25 (2020) 2000180.

WHO, Report of the WHO-China Joint Mission on coronavirus disease 2019 (COVID-19). 2020 (2020).

M. Poljak, M. Korva, N. K. Gasper, K. F. Komlos, M. Sagadin, T. Ursic, T. A. Zupanc, M. Petrovec, Clinical evaluation of the cobas SARS-CoV-2 test and a diagnostic platform switch during 48 hours in the midst of the COVID-19 pandemic, Journal of Clinical Microbiology 58 (2020).

E. Degli-Angeli, J. Dragavon, M. Huang, D. Lucic, G. Cloherty, K. R. Jerome, A. L. Greninger, R. W. Coombs, Validation and verification of the Abbott RealTime SARS-CoV-2 assay analytical and clinical performance, Journal of Clinical Virology (2020) 104474.

C. A. Hogan, M. K. Sahoo, C. Huang, N. Garamani, B. Stevens, J. Zehnder, B. A. Pinsky, Comparison of the Panther Fusion and a laboratory-developed test targeting the envelope gene for detection of SARS-CoV-2, Journal of Clinical Virology (2020) 104383.

W. Zhen, S. E, R. Manji, D. Schron, G. Berry, Clinical evaluation of three sample-to-answer platforms for detection of SARS-CoV-2, Journal of Clinical Microbiology 58 (2020).

C. M. Petrilli, S. A. Jones, J. Yang, H. Rajagopalan, L. F. O'Donnell, Y. Chernyak, K. Tobin, R. J. Cerfolio, F. Francois, L. I. Horwitz, Factors associated with hospitalization and critical illness among 4,103 patients with COVID-19 disease in New York City, MedRxiv (2020).

S. Garg, Hospitalization rates and characteristics of patients hospitalized with laboratory-confirmed coronavirus disease 2019–COVID-NET, 14 States, March 1-30, 2020, MMWR. Morbidity and mortality weekly report 69 (2020).

J. H. Beigel, K. M. Tomashek, L. E. Dodd, A. K. Mehta, B. S. Zingman, A. C. Kalil, E. Hohmann, H. Y. Chu, A. Luetkemeyer, S. Kline, D. Lopez de Castilla, Remdesivir for the treatment of Covid-19–preliminary report, New England Journal of Medicine (2020).

S. Richardson, J. S. Hirsch, M. Narasimhan, J. M. Crawford, T. McGinn, K. W. Davidson, D. P. Barnaby, L. B. Becker, J. D. Chelico, S. L. Cohen, J. Cookingham, Presenting characteristics, comorbidities, and outcomes among 5700 patients hospitalized with COVID-19 in the New York City area, Jama (2020).

ICNARC, ICNARC report on COVID-19 in critical care (2020).

M. G. Argenziano, S. L. Bruce, C. L. Slater, J. R. Tiao, M. R. Baldwin, R. G. Barr, B. P. Chang, K. H. Chau, J. J. Choi, N. Gavin, P. Goyal, Characterization and clinical course of 1000 patients with coronavirus disease 2019 in New York: retrospective case series, bmj 369 (2020).
